# Enteric pathogen burden and co-infection patterns across age and a rural-urban gradient: findings from the ECoMiD birth cohort, Northern Ecuador

**DOI:** 10.64898/2026.07.08.26357325

**Authors:** Nicolette A. Zhou, Caitlin Hemlock, Kelsey J. Jesser, Christine S. Fagnant-Sperati, Jesse D. Contreras, Benjamin F. Arnold, William Cevallos, Gabriel Trueba, Gwenyth O. Lee, Joseph N.S. Eisenberg, Karen Levy, the ECoMiD Investigators

## Abstract

Enteric pathogen infections are a major global health challenge, influenced by a variety of host and environmental factors, and their clinical presentation and treatment can be complicated by the presence of co-infections. The prevalence of enteric infections and co-infections tend to vary between rural and urban contexts, likely driven by underlying environmental, geographic, and demographic characteristics. To improve understanding of urbanicity and age on enteric pathogen prevalence and on co-infection risk, we measured 22 enteric pathogens in fecal samples collected from children aged 6, 12, and 18 months across a rural-urban gradient within the ECoMiD birth cohort study (*n*=473). Enteric pathogen burden was high and increased with age, with at least one pathogen detected in 91% of children at 6 months, 97% at 12 months, and 98% at 18 months. However, prevalence of some pathogens— notably *Salmonella enterica,* enterovirus, and rotavirus— decreased with age. Co-infections were also common (88%), and children were infected with as many as 11 pathogens simultaneously. The most frequently observed co-infection profiles included enteroaggregative *E. coli* and atypical enteropathogenic *E. coli*, followed by combinations with diffusely adherent *E. coli*, enterovirus, enterotoxigenic *E. coli*, and/or adenovirus. Enteric pathogen detection generally was higher in more rural settings, though patterns varied by pathogen. These results provide useful information for future examination of pathogen dynamics of co-occurrence. Given the ubiquity of enteric infections in high transmission settings, strategies that aim to reduce overall microbial exposure may be needed to supplement interventions targeting control of individual pathogens.

**AUTHOR SUMMARY:** Children living in low-resource settings are frequently exposed to pathogens that cause enteric infections, which can contribute to diarrhea, poor growth, and long-term health problems. Many children are infected with more than one pathogen at the same time, but we still know relatively little about how these infections occur together or how they vary across different environments. We examined stool samples collected from children during their first 18 months of life in communities ranging from urban to rural areas of northern Ecuador. We found that nearly every child carried at least one pathogen, and most carried several simultaneously. The number of pathogens increased as children grew older, and children living in more rural communities generally had a higher burden of infection. We also found that some pathogens occurred together more often than expected by chance, suggesting they may share common transmission routes or interact in ways that deserve further study. Our findings show that children in high-transmission settings experience a substantial burden of multiple simultaneous infections from an early age. These results support the need for interventions that reduce overall exposure to enteric pathogens, alongside strategies that target individual pathogens such as vaccines.

## INTRODUCTION

Enteric pathogen infections remain a major global health challenge, particularly affecting children under five years of age in low- and middle-income countries (LMICs)(1,2). These infections are caused by a diverse array of bacterial, viral, and parasitic pathogens that are transmitted fecal-orally and colonize or infect the gastrointestinal tract, resulting in diarrhea, subclinical gut dysfunction, malnutrition, and long-term developmental impacts(3). Advances in molecular diagnostic tools have greatly improved the ease with which multiple pathogens can be simultaneously detected within a single host. These tools have revealed high pathogen burdens and rates of co-infection globally, particularly in children in LMICs where unimproved water, sanitation, and hygiene (WASH) facilitates transmission of enteric pathogens(4,5).

Whether symptomatic with acute diarrhea or asymptomatic(4,6–8), these co-infections can complicate clinical presentation and diagnosis and may influence disease severity, pathogen shedding dynamics, immune responses, and response to treatment(9–11). In theory, pathogen-pathogen interactions may be synergistic or antagonistic, shaping infection persistence and downstream health consequences in ways that differ when pathogens are present in isolation. However, these interactions are not well characterized, including the types of pathogens most frequently involved in co-infections.

Beyond clinical implications, the presence of co-infections complicates questions of etiology; i.e., which pathogens cause disease. Ambiguous etiology has implications for estimating morbidity and mortality attributable to any particular pathogen, and ultimately for prioritization of pathogens for new interventions and trial study design. For example, accounting for co-infections led to an 11.3% increase in the efficacy estimate of a rotavirus vaccine trial(12), as vaccines may seem less effective if other co-infections cause symptoms in individuals who receive the vaccine. Assessing co-infections across multiple enteric pathogens can provide a more nuanced understanding of an individual’s overall pathogen burden. However, enteric pathogen transmission is a complex process influenced by a myriad of factors, such as socio-economic status, the environment, and geography. Thus, population-level studies are needed to assess variation in pathogen burden across populations and to identify epidemiological patterns and environmental, geographic, and age-specific risk factors for co-infections, and interactive effects of these multiple co-occurring infections (11,13–15).

Urbanicity has been associated with differences in pathogen prevalence and diversity across settings, reflecting variations in infrastructure, population density, sanitation, and healthcare access(16), though this relationship is complex. Urban areas may benefit from better access to improved water and sanitation infrastructure compared to more rural areas; however, higher population density and higher rates of movement can increase transmission risk(17–21). Conversely, rural areas often face challenges of uncontained environmental contamination and limited healthcare access but may have lower person-to-person transmission due to dispersed populations(20–22). While several multisite studies have examined differences in enteric pathogen prevalence between countries(1,6), examining sites across an urban-rural gradient within the same country can help isolate localized geographic and environmental exposure variability within a region that is otherwise relatively similar in terms of cultural, social and genetic factors.

Age is another important variable that additionally influences enteric infection risk, particularly within the first two years of life, when susceptibility changes rapidly as children transition from maternal protection to developing their own immune responses. Infants and young children exhibit higher susceptibility to enteric infections, partly due to this immature immune response and increased exposure through exploratory behaviors and changing diets(3). Early childhood is a critical period for gut microbiome development, which can affect susceptibility to both single and multiple infections(23). Understanding how the overall burden of enteric pathogen infections and co-infections is influenced by age can guide age-specific interventions and clinical management.

In this study, we examined the prevalence of 22 individual enteric pathogens in children aged 6, 12, and 18 months old across a rural-urban gradient from the longitudinal ‘Enteropatógenos, Crecimiento, Microbioma, y Diarrea’ (ECoMiD) study(24). We assessed how individual pathogen carriage, co-infection prevalence, and overall infection burden varied by urbanicity and age. Evaluating individual pathogens provides insight into their carriage and transmission patterns across a range of settings, while examining co-infections helps capture the broader cumulative burden of infection and exposure. These analyses provide valuable information for future holistic strategies to improve child health.

## METHODS

### Study design

Participants were recruited from ten ECoMiD study communities in Northern coastal Ecuador, grouped into four categories: i) Urban: the city of Esmeraldas, the provincial capital (population ∼162,000), ii) Intermediate: the town of Borbón, the commercial center of a politically defined rural region (population ∼5,000), iii) Rural road: rural villages that are accessible by road near Borbón, including the communities of Timbiré, Selva Alegre, Colón Eloy, and Maldonado (populations ∼500-1,000), and iv) Rural river: rural villages that are only accessible by river or with limited car accessibility, including Santo Domingo, Zancudo, Colón de Onzole, and San Francisco (populations ∼200–700) (Figure 1). Additional details about the ECoMiD study have been published previously(24).

**Figure 1.**
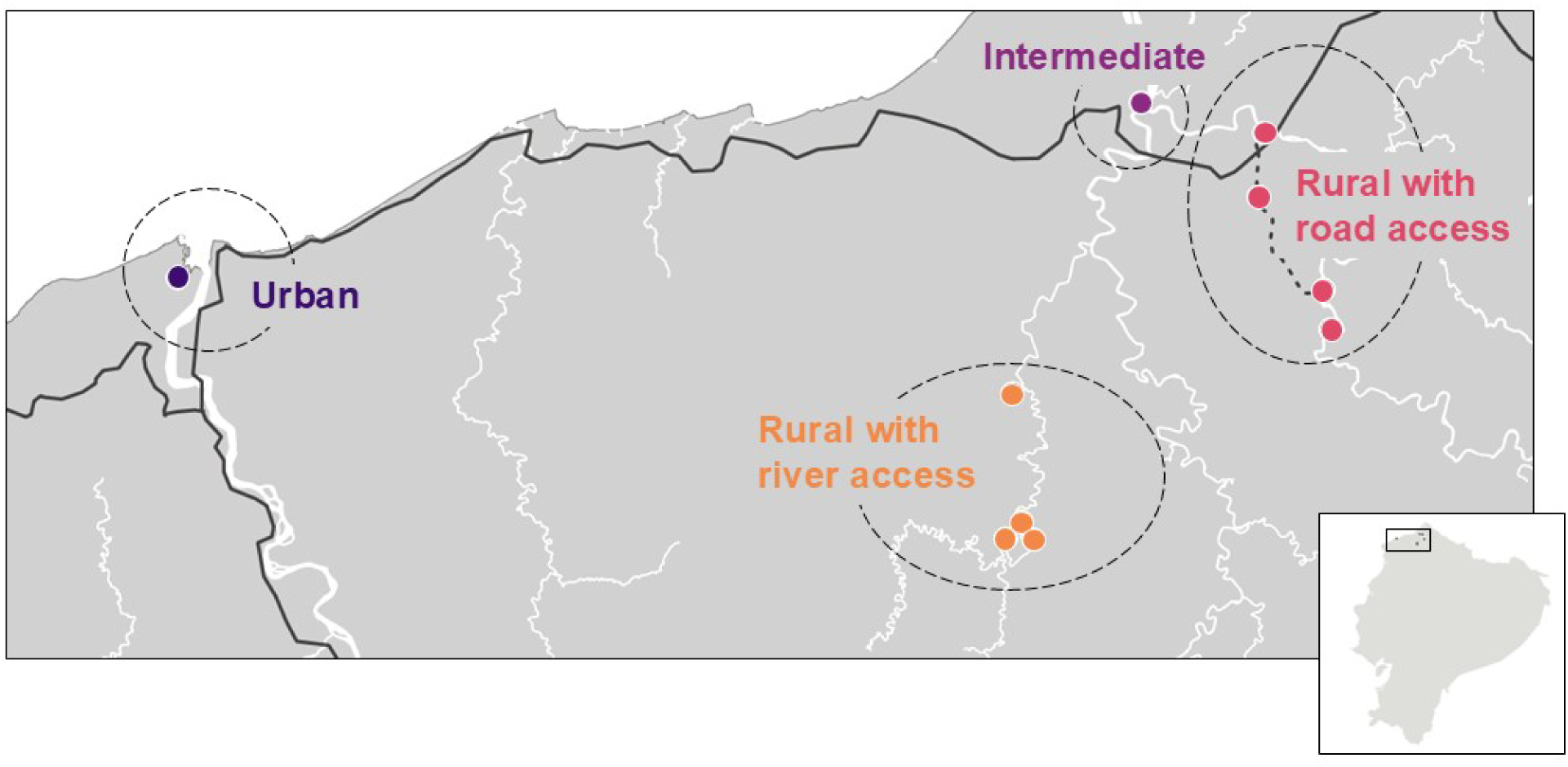
Map of the study communities in the ECoMiD study in Northern coastal Ecuador, including an urban site (Esmeraldas), intermediate site (Borbón), four rural road communities (Maldonado, Timbiré, Selva Alegre, Colón Eloy), and four rural river communities (Colón de Onzole, Santo Domingo, Zancudo, San Francisco).

The study protocol was approved by the institutional review boards of the University of Washington (UW; IRB STUDY00014270), Emory University (IRB00101202), and the Universidad San Francisco de Quito (USFQ; 2018–022M), as well as the Ministry of Health of Ecuador (MSPCURI000253-4).

### Household characteristics

Surveys were conducted by trained local fieldworkers to collect data about mother’s demographics (education and occupation), household socioeconomic status (SES), water, sanitation, and hygiene (WASH) infrastructure, and animal exposure, as previously described(24,25).

### Stool sample collection and nucleic acid extraction

Child stool samples were collected, as previously described,(25) at 6, 12, and 18 months of age (±6 weeks). Briefly, caregivers placed child stool samples in sterile containers and placed them in a refrigerator or study-provided cooler. In the field lab, samples were aliquoted and stored in liquid nitrogen dewars(24). Stool aliquots were transported to USFQ where total nucleic acids (TNA) were stored at − 80°C prior to and following extraction using the QIAamp Fast DNA Stool Mini Kit (Qiagen, Germantown, MD). Modifications of the kit extraction protocol included addition of 370 mg Sigma glass beads, acid-washed 212-300 μM (50-70 US Sieve), 5 μL Phocine Herpes Virus 1 (PhHV) stock as a DNA extraction control, 1 μL Bacteriophage MS2 stock as an RNA extraction control, and 10 μL ZymoBIOMICS Spike-in Control as a next generation sequencing control with the InhibitEX buffer.

Samples were bead-beaten for 1 minute (FastPrep-24 [MP Biomedicals, Irvine, CA]) and placed on ice for 1 minute 3 times to ensure full lysis. Finally, the samples were incubated for 5 minutes at 95°C, and the manufacturing protocol resumed. Frozen TNA samples were shipped from USFQ to University of Washington (UW) on Techni-Ice (Frankston, Australia) and stored at −80°C until analyzed.

### TaqMan array card analysis

Extracted TNA were analyzed for bacterial, viral, parasite, and antimicrobial resistance gene targets using TaqMan Array Cards (TAC), as previously described.(25) Briefly, TAC controls included PhHV (DNA control seeded during extraction), MS2 (RNA control seeded during extraction), and the pan *E. coli* gene target *uidA* (expected to be present in stool samples). Positive controls consisted of four custom plasmids, and a 10-fold serial dilution of these controls was run every 20 cards. A no-template control was included on each card. Samples and controls were analyzed using the AgPath-ID One-Step RT-PCR Reagents (ThermoFisher Scientific, Waltham, MA) on a QuantStudio 7 Flex instrument (ThermoFisher Scientific), with the following cycling conditions: 45°C for 20 minutes, 95°C for 10 minutes, then 40 cycles of 95°C for 15 seconds and 60°C for 1 minute. Results for individual assays were obtained as quantal data with Ct values <35 considered positive. The presence or absence of each enteric pathogen was determined as previously reported and as shown in Table S1(25). Extraction controls were analyzed using TAC and if a target was positive, samples for that target from that extraction batch were considered missing if the Ct value was equal to or greater than the extraction control Ct value. This occurred in 77 of 42,804 assays for pathogen targets included in this study.

### Outcomes

Outcomes of interest included any infection, number of distinct pathogens, infection with any bacteria, infection with any virus, infection with any parasite, and infection with individual pathogens. Infection in this study refers to detection by (RT)-qPCR. Outcome data were treated as binary presence/absence except for number of pathogens, which was treated as count data. Individual pathogen outcomes included ten bacteria: diffusely adherent *E. coli* (DAEC), enteroaggregative *E. coli* (EAEC), shigatoxigenic *E. coli* (STEC), enterohemorrhagic *E. coli* (EHEC), enteroinvasive *E. coli* (EIEC)/*Shigella*, enterotoxigenic *E. coli* (ETEC), atypical enteropathogenic *E. coli* (aEPEC), typical enteropathogenic *E. coli* (tEPEC), *Campylobacter* spp., and *Salmonella enterica* (non-typhoidal and typhoidal *Salmonella* targets were included); six viruses: adenovirus, astrovirus, enterovirus, norovirus, rotavirus, and sapovirus; and six parasites: *Ascaris lumbricoides*, *Cryptosporidium* spp., *Cyclospora cayetanensis, Entamoeba histolytica, Giardia lamblia*, and *Trichuris trichiura*.

Sensitivity analyses were conducted for EAEC and enterovirus. For EAEC, it was excluded from the ‘any infection’, ‘pathogen number’, and ‘any bacteria’ outcomes. This was completed due to the high prevalence of EAEC, uncertainty about its status as a pathogen, and potential cross reactivity of gene targets(26,27). Enterovirus was excluded from the ‘any virus’ outcome due to its high prevalence relative to other viruses in this study and its clinical and epidemiological profile. While enteroviruses are important pathogens causing a range of serious illnesses, they are often associated with brief enteric illnesses(28).

### Statistical analyses

To assess the association between age, community type, and enteric pathogen infection, we fit modified Poisson models using generalized estimating equations (GEE) to estimate prevalence ratios (or count ratios for count outcomes), accounting for child-level clustering with robust standard errors. We adjusted for birth year to control for cohort effects (and thus time effects due to the inclusion of age in the model), and the season during which the sample was collected (rainy [January-May] or dry [June-December]). For any models that did not converge, we switched to a binomial distribution, resulting in Odds Ratios (ORs) as the estimated parameter. For individual pathogen infection outcomes, we estimated associations only for outcomes with prevalence >1%. We used a complete case analysis approach for all models.

Pairwise co-occurrence analyses were conducted using the *cooccur* package in R. This approach compares the frequency that pairs of pathogens were detected simultaneously compared to the expected frequency if the two pathogens were independent. ‘Positive’ indicates that the two organisms were detected simultaneously more often than expected by chance; ‘Random’ indicates that detection of the two organisms did not differ significantly from the random null hypothesis; and ‘Negative’ indicates that the two organisms were detected simultaneously less often than expected by chance. This method factors in the underlying prevalence of the individual pathogens when calculating results.

All analyses were conducted in R v4.3.3.

## RESULTS

### Study sample

A total of 521 children were enrolled in the ECoMiD cohort between 2019-2022. TAC analysis of stool samples collected at ages 6, 12, and/or 18 months was completed for 476 unique participants, with a total of 1181 stool samples analyzed. Data are not available for 45 mother-child dyads as they left the study for the following reasons: moved (27), withdrew from the study (7), child passed away (4), lost contact with the family (3), unable to maintain contact due to social unrest in Esmeraldas (3), and caregiver declined to consent to stool collections (1). The majority of samples were collected within six weeks of the target age (Figure S1), and excluding those that were collected outside this window resulted in a total of 1,154 samples, including samples from children aged 6 (*n*=359), 12 (*n*=404), and 18 (*n*=391) months (473 unique children total). Samples were distributed across the urban site (*n*=265), intermediate site (*n*=349), rural road communities (*n*=420), and rural river communities (*n*=120).

Household environmental conditions varied by community type (Figure 2). At enrollment, urban households were more likely to have piped water, be of higher SES, and have a higher level of maternal education than households in intermediate and rural communities. Households in urban and rural road communities showed similar trends, with higher levels of improved sanitation and a higher level of animal ownership compared to intermediate and rural river communities, while mothers having a job outside of the home was low across all communities (<20%).

**Figure 2.**
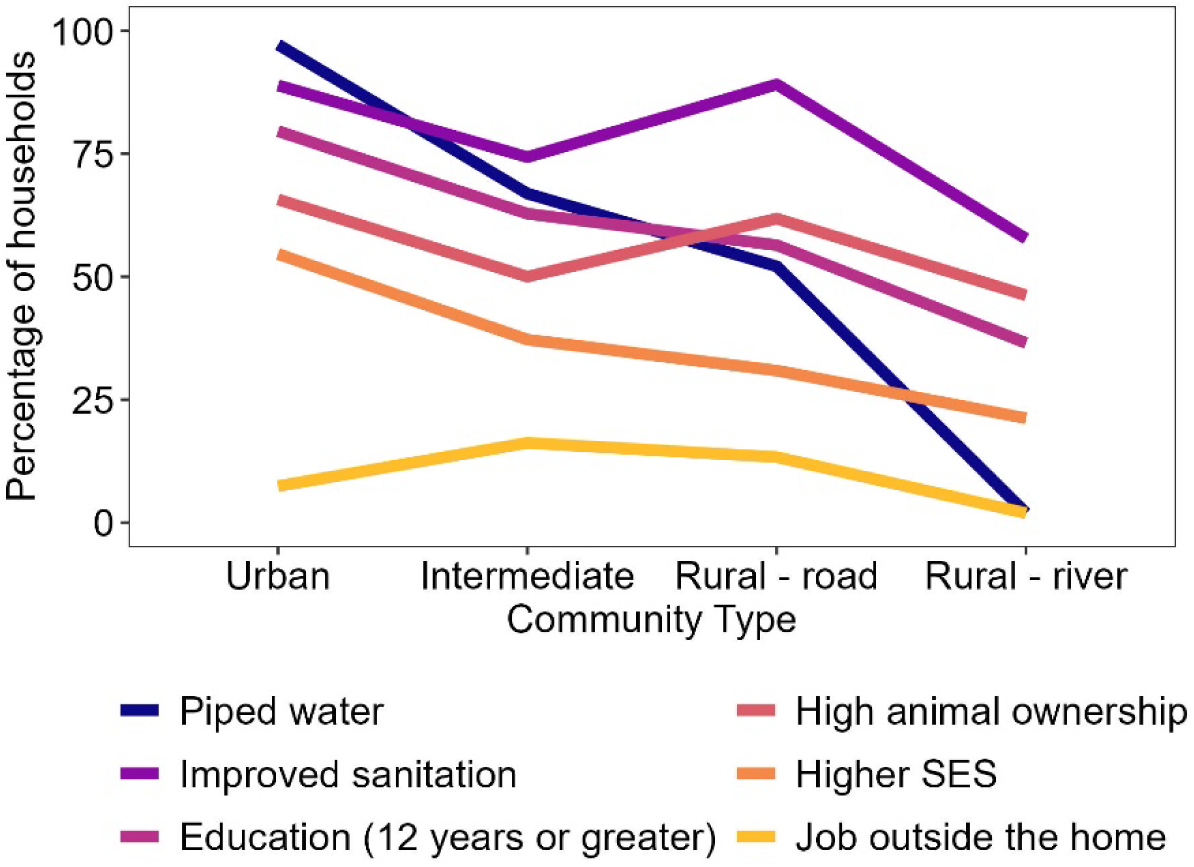
Household environmental conditions and socioeconomic status (SES) by community type. SES and animal exposure scores were determined as previously described with high animal ownership having a score greater than 2 out of 4, and higher SES was within the upper 2 quintiles.

### Enteric pathogen prevalence

Enteric pathogen detection was high overall, with 95% of children positive for at least one pathogen (Figure 3, Table S1). Bacterial pathogens were highly prevalent (93%); EAEC was the most highly prevalent bacterium across all ages and sites (73%). The prevalence of bacterial pathogens remained high when we excluded EAEC (84%). Other frequently detected diarrheagenic *E. coli* pathotypes were: DAEC (46%), aEPEC (44%), and ETEC (32%). The remaining six bacterial pathogens analyzed (STEC, tEPEC, EHEC, EIEC/*Shigella*, *Campylobacter* spp., and *Salmonella enterica*) were present at rates of 1.6-21%. Viral pathogens were detected in 61% of children. Enteroviruses were the most frequently detected virus (36% overall), followed by adenovirus (32%) and norovirus (11%). Rotavirus, sapovirus, and astrovirus were less common across all settings (3–5%). Parasite infections were less prevalent overall (30%). *Giardia lamblia* was the most common parasite (19%), followed by *Cryptosporidium* spp. (8.6%) and *Ascaris lumbricoides* (7.9%). Other parasites, including *Trichuris trichiura*, *Cyclospora cayetanensis*, and *Entamoeba histolytica*, were detected at low frequencies (<2%).

**Figure 3.**
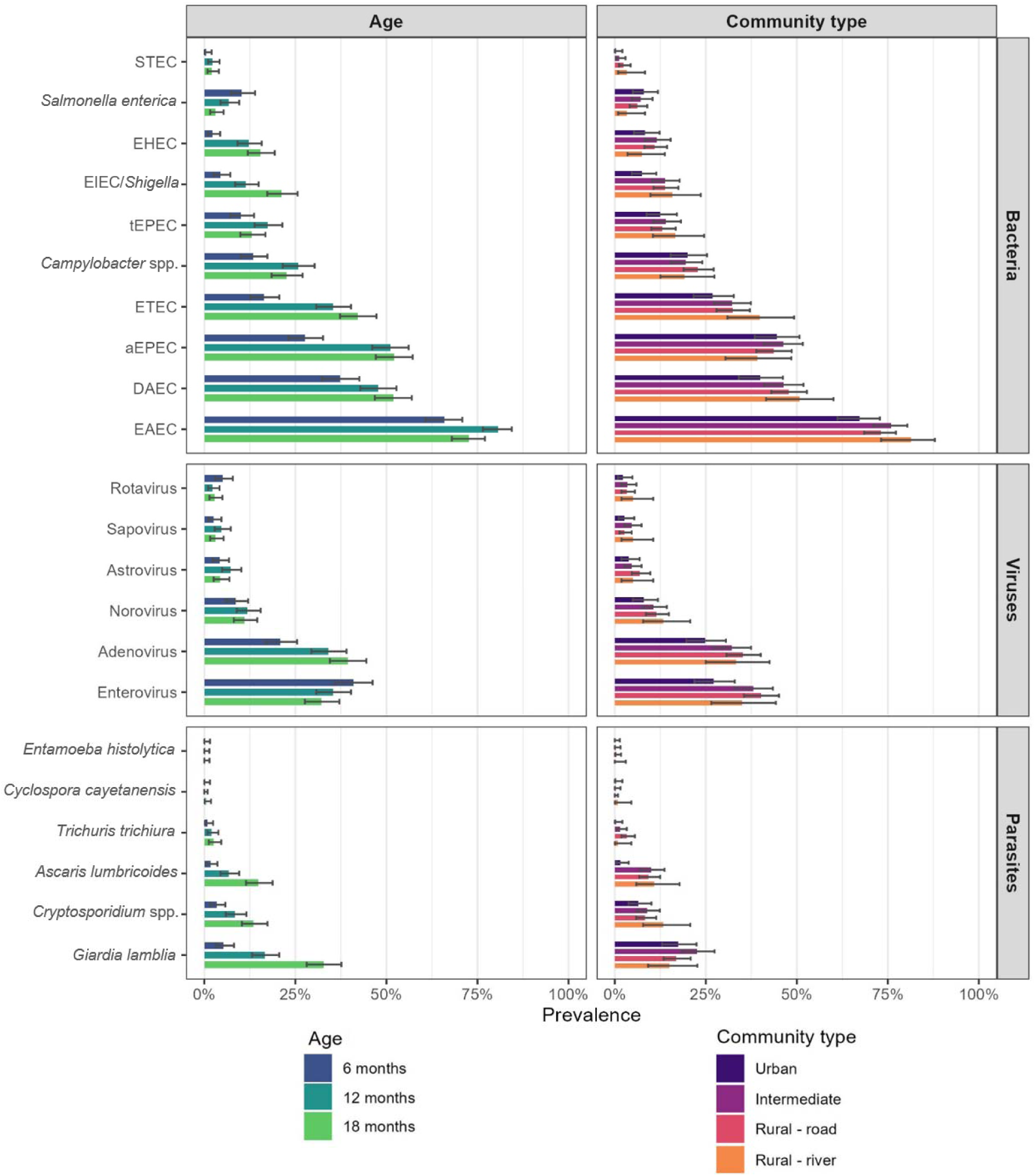
Prevalence of bacteria, virus, and parasite pathogens in child stool samples by age (±6 weeks) (*n*=359 at 6 months, *n*=404 at 12 months, and *n*=391 at 18 months) and community type (*n*=265 in urban, *n*=349 in intermediate, *n*=420 in rural road, and *n*=120 in rural river). Error bars show the upper and lower confidence intervals around the mean. Where aEPEC is atypical enteropathogenic *E. coli*, DAEC is diffusely adherent *E. coli*, EAEC is enteroaggregative *E. coli*, EHEC is enterohemorrhagic *E. coli*, EIEC is enteroinvasive *E. coli*, ETEC is enterotoxigenic *E. coli*, STEC is shigatoxigenic *E. coli*, and tEPEC is typical enteropathogenic *E. coli*.

Co-infections were common, with 88% of children positive for multiple pathogens and 79% positive for multiple pathogens when excluding EAEC (Table S1). Children commonly carried between two and five pathogens (median: 4, interquartile range [IQR]: 2–5) and were infected with up to 11 pathogens simultaneously. There were 659 unique co-infection profiles (Table S2), and the most frequently observed profile was EAEC and aEPEC (n=23 [2%] samples), followed by combinations with DAEC, enterovirus, ETEC, and/or adenovirus (Figure 4). Viral–bacterial profiles were the most frequently observed (40.5% of profiles and 42.9% of samples with co-infections), followed by bacterial-viral-parasitic co-infections (34.3% of profiles and 24.3% of samples with co-infections), bacterial-bacterial (13.4% of profiles and 22.4% of samples with co-infections), and bacterial–parasitic (10.6% of profiles and 9.3% of samples with co-infections). Frequently observed co-infection profiles (>5 samples with the same profile) typically contained two or three pathogens, with two exceptions containing four pathogens.

**Figure 4.**
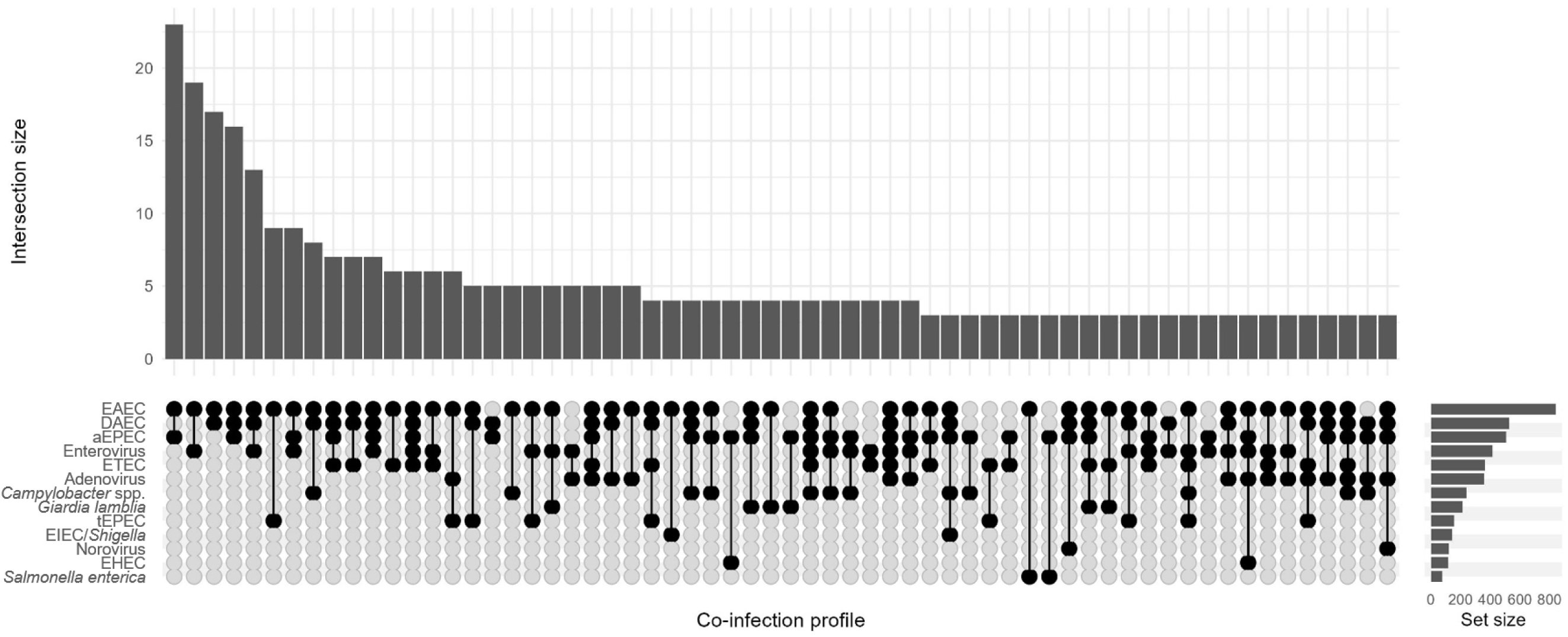
Co-infection profiles for individual child stool samples collected at 6, 12, and 18 months (±6 weeks). Profiles present in three or more samples are shown. Samples with one pathogen or less detected are not included. Intersection size is the number of samples that have the same co-infection profile. Set size is the number of samples that are positive for an individual pathogen. Where aEPEC is atypical enteropathogenic *E. coli*, DAEC is diffusely adherent *E. coli*, EAEC is enteroaggregative *E. coli*, EHEC is enterohemorrhagic *E. coli,* EIEC is enteroinvasive *E. coli*, ETEC is enterotoxigenic *E. coli*, and tEPEC is typical enteropathogenic *E. coli*.

Pairwise co-occurrence analysis showed non-random clustering of pathogens (Figure 5). The majority of significant co-occurrence was positive, indicating that the pathogens co-occurred more often than would be expected by chance. Positive significant co-occurrence most frequently occurred among combinations involving adenovirus, ETEC, *Giardia lamblia*, EIEC/*Shigella*, *Campylobacter* spp., DAEC, and EHEC (Figure 5).

**Figure 5.**
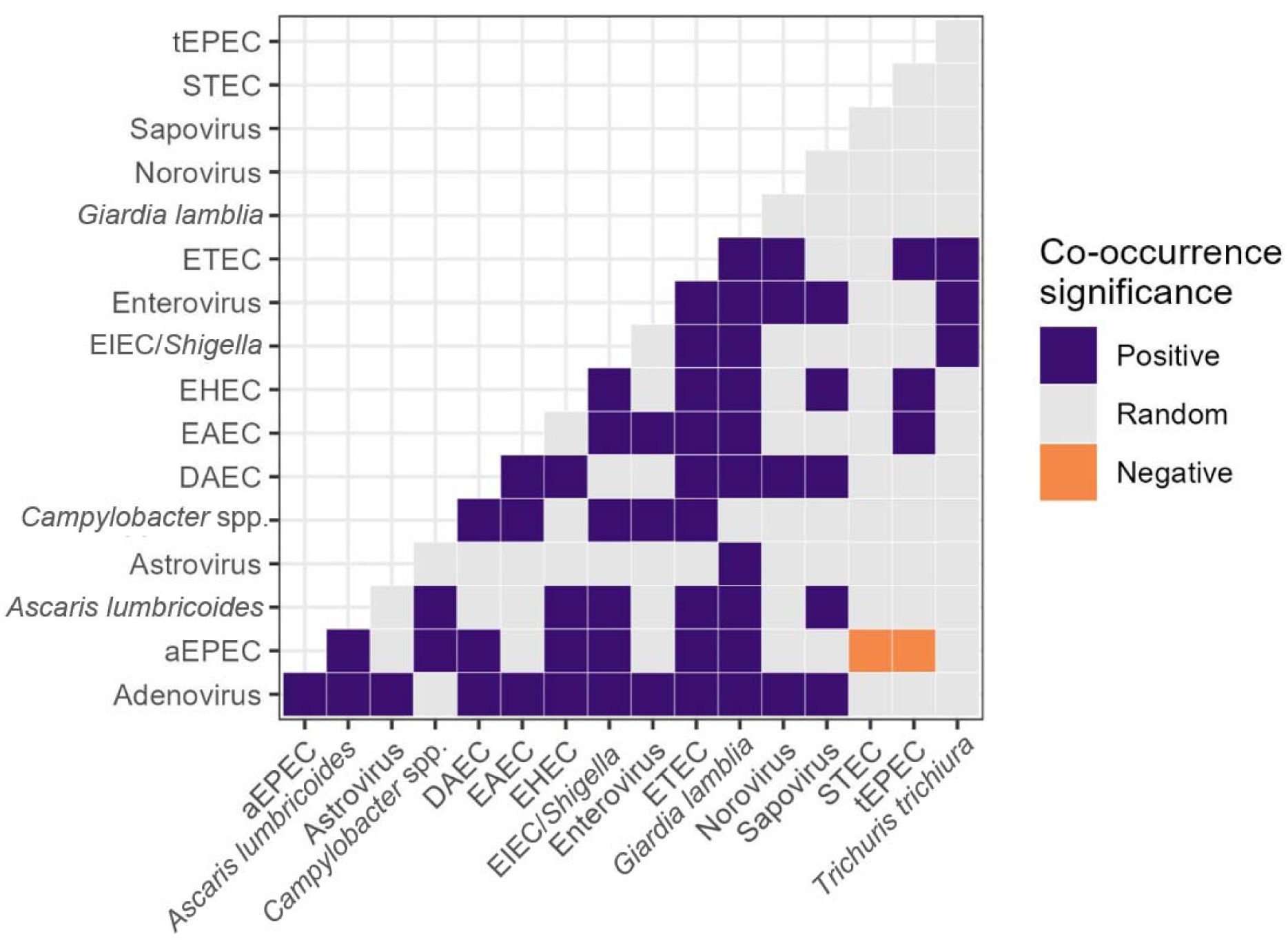
Pathogen co-occurrence heatmap. ‘Positive’ indicates that the two organisms were detected simultaneously more often than expected by chance; ‘Random’ indicates that detection of the two organisms did not differ significantly from the random null hypothesis; and ‘Negative’ indicates that the two organisms were detected simultaneously less often than expected by chance. The negative associations seen between aEPEC, tEPEC, and STEC are due to how these organisms are defined. Where aEPEC is atypical enteropathogenic *E. coli*, DAEC is diffusely adherent *E. coli*, EAEC is enteroaggregative *E. coli*, EHEC is enterohemorrhagic *E. coli*, EIEC is enteroinvasive *E. coli*, ETEC is enterotoxigenic *E. coli*, STEC is shigatoxigenic *E. coli*, and tEPEC is typical enteropathogenic *E. coli*.

### Infection patterns by age

Enteric pathogen infection generally increased with child age, with some exceptions (Figure 3, Table S1). At 6 months, 91% of children were positive for at least one pathogen, rising to 97% at 12 months and 98% at 18 months.

For bacterial pathogens, prevalence of any bacteria increased with age, with detection of at least one in 84% of children at 6 months, 96% at 12 months, and 97% at 18 months old (Figure 3). Compared to 6-month-old children, 12-month-olds had a 13% increased prevalence of any bacterial infection (95% CI: 8-19%) and 18-month-olds had a 15% increased prevalence (95% CI: 9-20%) (Figure 6, Table S3). This increase in infection prevalence with age was also seen for individual pathogens at both 12 and 18 months for DAEC, EHEC, EIEC/*Shigella*, aEPEC, and *Campylobacter* spp., and at 12 months for EAEC and tEPEC. The exception was *Salmonella enterica*, which had a 72% decreased prevalence (95% CI: 47-85%) at 18 months. Due to the aforementioned limitations for EAEC, we excluded EAEC from the ‘any infection’, ‘pathogen number’, and ‘any bacteria’ outcomes and saw minimal impact on their associations with age with no change in statistical significance (Figure S3).

**Figure 6.**
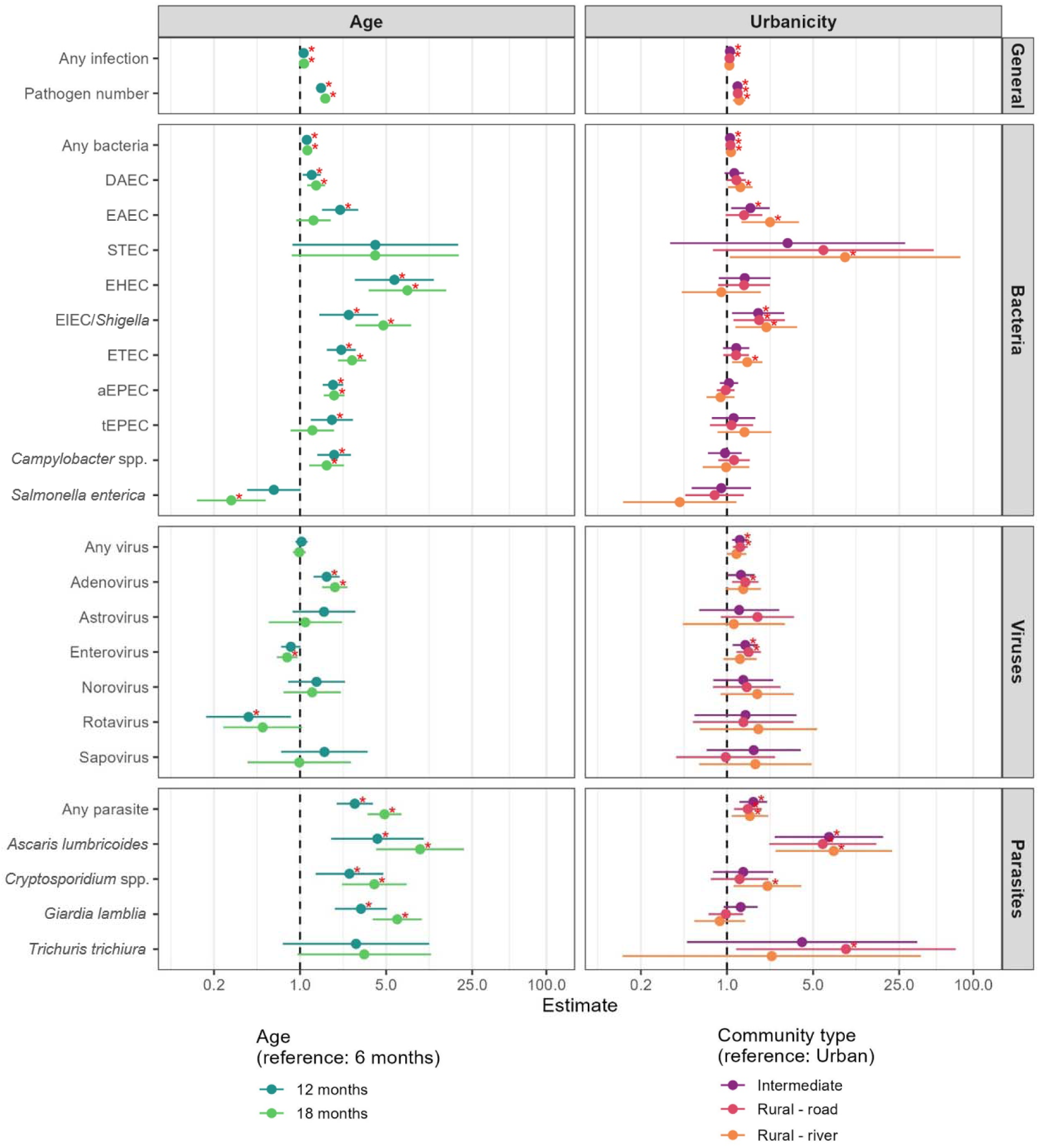
Associations between age (±6 weeks), urbanicity, and pathogen infection. All estimates are adjusted for the child’s birth year and season. Age models are adjusted for urbanicity and urbanicity models are adjusted for age. The *x*-axes are a count ratio for number of pathogens, odds ratio for EAEC and norovirus, and a prevalence ratio for all other outcomes. Asterisks indicate significance at *p*<0.05.

Viral enteric pathogens did not show the same overall upward trend with age. We detected viruses in ≥60% of children at 6, 12, and 18 months (Figure 3). However, trends varied by individual viral pathogens. Adenovirus increased with age (21% [6 months] to 39% [18 months]), with a 64% higher prevalence at 12 months (95% CI: 29-109%) and 92% increase at 18 months (95% CI: 51-143%) when compared to 6 months (Figure 6, Table S3). In contrast, enterovirus prevalence decreased with age (41% [6 months] to 32% [18 months]) and showed a 21% decreased prevalence (95% CI: 5-35%) at 18 months when compared to 6 months. Due to the high prevalence of enterovirus compared to other viruses, we examined the ‘any virus’ outcome with enterovirus excluded; we saw a shift with increasing age, from no difference by age with enterovirus included, to an increased prevalence of any virus infection with age with enterovirus excluded (Figure S3). Rotavirus also showed a 62% decreased prevalence (95% CI: 16-83%) at 12 months when compared to 6 months.

Parasite prevalence increased with age (10% of children at 6 months, 28% at 12 months, and 50% at 18 months; Figure 3), mirroring the pattern observed for bacterial pathogens. Compared to 6-month-olds, 12-month-olds were associated with 178% increased prevalence of any parasite infection (95% CI: 98-291%) and 18-month-olds were associated with 385% increased prevalence of any parasite infection (95% CI: 255-567%) (Figure 6, Table S3). This trend was seen for individual pathogens as well, including *Ascaris lumbricoides, Cryptosporidium* spp., and *Giardia lamblia*.

Co-infections were common across all age groups and increased with age: 76% of 6-month-olds had more than one pathogen, compared to 92% of 12-month-olds and 94% of 18-month-olds. Similarly, the total count of pathogens per child increased with age (Table S3), with the range increasing from 2.3-3.2 at 6 months, to 3.7-4.4 at 12 months, and 3.9-4.8 at 18 months (Figure 7).

**Figure 7.**
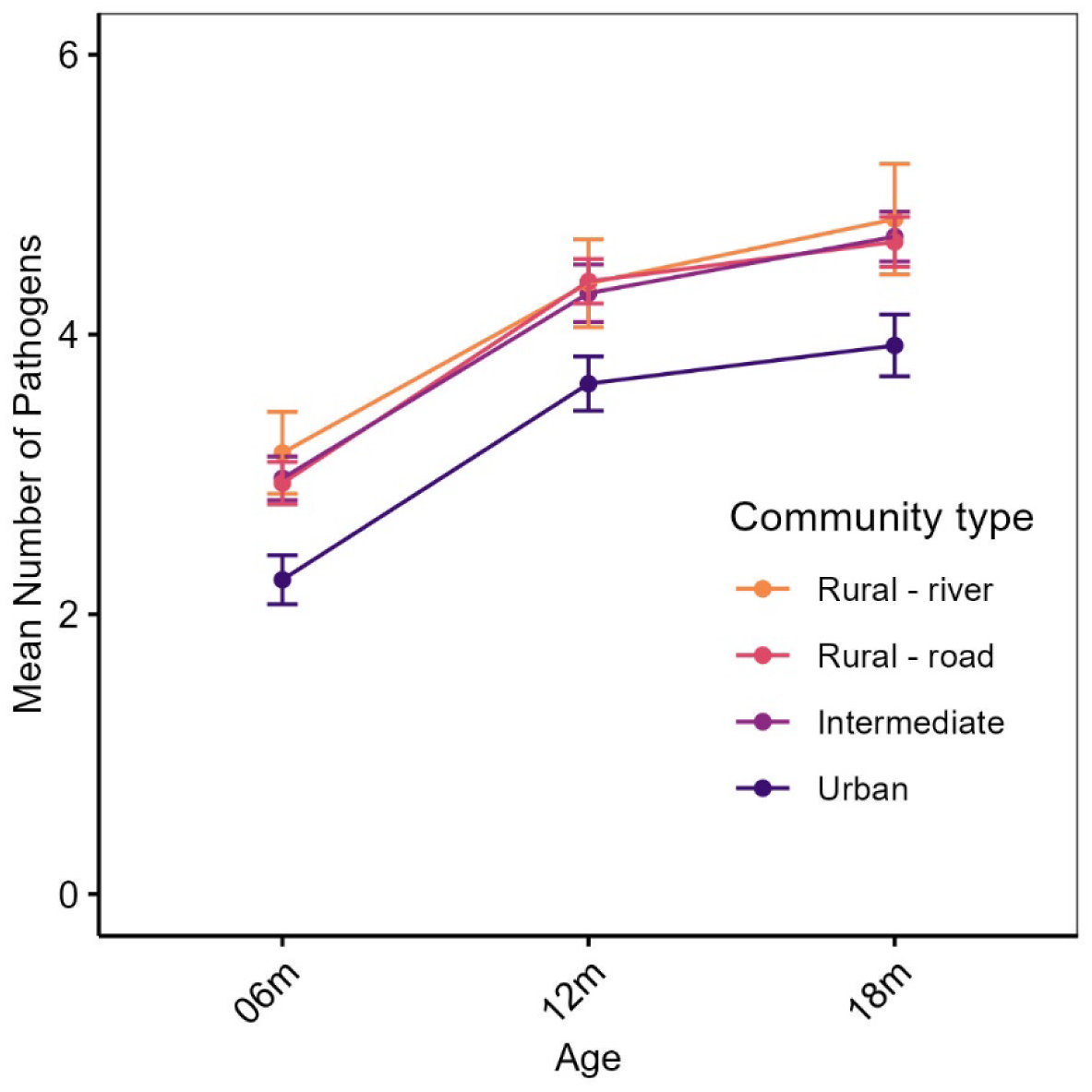
Mean number of pathogens detected per child at 6, 12, and 18 months (±6 weeks) by community type with error bars representing standard error.

There were no differences in results when the analytic sample included all samples (i.e. was not limited to children sampled in the target window of ±6 weeks only), including for any bacteria, any virus, any parasite, or co-infections (Figure S2).

### Infection patterns by urbanicity

Enteric pathogen detection was the lowest in the urban site (Figure 3, Table S1), with prevalence of any pathogen in 92% of children in the urban site, compared to 97%, 96%, and 96% at intermediate, rural road, and rural river communities, respectively. Compared to the urban site, living in the intermediate and rural communities yielded a significant increased prevalence of any bacterial, viral, or parasite infection with the exception of any viral infection in the rural river communities (Figure 6 and Table S3).

This trend of lower enteric pathogen infections in the urban site was also seen for individual pathogens (Figure 6 and Table S3). Living in the intermediate site was associated with an increased prevalence of EAEC and enterovirus infection, living in the rural road communities was associated with an increased prevalence of adenovirus, enterovirus, and *Trichuris trichiura* infection, and living in the rural river communities was associated with an increased prevalence of EAEC, DAEC, STEC, ETEC, and *Cryptosporidium* spp. infection, compared to living in the urban site. Finally, the prevalence of EIEC/*Shigella* and *Ascaris lumbricoides* were higher among children living in all communities compared to those living in the urban site. Due to the high prevalence of EAEC and enterovirus compared to other bacteria and viruses, we examined excluding EAEC from the ‘any infection’, ‘pathogen number’, and ‘any bacteria’ outcomes and enterovirus from the ‘any virus’ outcome and saw minimal impact on their associations with community type (Figure S3). Excluding EAEC increased the confidence intervals around the estimated effect sizes for any bacterial infection in the intermediate site compared to the urban site, eliminating statistical significance. Excluding enterovirus increased the estimated effect sizes for any viral infection in the rural river communities compared to the urban site, resulting in statistical significance.

Co-infections were common across all age groups and communities but were more frequently detected in more remote communities (Table S3). Children living in the urban community consistently exhibited lower mean numbers of pathogens (Figure 7; results for all children not limited to ±6 weeks are shown in Figure S4). At 6 months of age, the mean number of pathogens detected in the urban site was 2.3, compared to 3.2 in rural river communities. This trend persisted across all measured age groups.

## DISCUSSION

Our findings reveal a high burden of enteric pathogen carriage among children aged 6 to 18 months in Northern Ecuador across an urban-rural gradient. The high rate of co-infections highlights the importance of examining pathogen infections in combination, given their potential synergistic effects on child health and development(9–11). Although infection risk was generally higher in the rural and intermediate communities, reflecting the differences in environmental exposure conditions and WASH access, the burden of infection among children in the urban site was also high, particularly when compared to urban settings in high-income countries(29,30). This suggests that even substantial improvements in infrastructure such as piped water and improved sanitation (Figure 2) are insufficient to eliminate the persistent background burden of infection. The age-related trends we found further underscore the cumulative nature of exposure. These findings suggest that in areas with high enteric pathogen transmission, strategies that aim to reduce overall microbial exposure may be needed to supplement interventions targeting control of individual pathogens.

Our findings for the enteric pathogen profiles in this cohort reflect a setting with sustained exposure to multiple clinically relevant pathogens, rather than dominance by a single organism. Of the thirteen pathogens examined by the recent Global Burden of Disease (GBD) Study(31), three (*Vibrio cholerae, Aeromonas* spp., and *Clostridioides difficile*) were not measured in this study, while the remaining ten were detected at varying overall prevalences (<10% for four pathogens and 11-32% for six pathogens; Table S1). The GBD Study estimated that, among children under five years of age, rotavirus, *Shigella* spp., and adenovirus were leading contributors to diarrhea disability-adjusted life-years (DALYs) and deaths, with additional pathogens, including *Cryptosporidium* spp., tEPEC, ETEC, norovirus, and *Campylobacter* spp., also exceeding the 5% fatal population attributable fraction (PAF) threshold.

Rotavirus prevalence was low in the ECoMiD cohort (3.3%), likely reflecting the impact of vaccination, which differs from its historically high contribution to global diarrheal burden and highlights the importance of continued surveillance and local epidemiologic context when interpreting pathogen prevalence. In contrast, the high prevalence of adenovirus (32%) and ETEC (32%) across ages and settings in our cohort suggest widespread and persistent transmission rather than localized outbreaks. Similarly, while *Shigella* (13%) and *Cryptosporidium* spp. (8.6%) were detected at more modest prevalences, their disproportionate contribution to severe disease and mortality suggests that prevalence alone underestimates their importance. Finally, the frequent detection of pathogens with lower global DALY contributions, such as *Campylobacter* spp. (21%) and *Salmonella enterica* (6.6%), points to a complex environment in which multiple co-circulating pathogens may influence child health outcomes. This underscores the need to interpret individual pathogen prevalence within broader contexts, whether global burden frameworks or local geographical and environmental contexts.

Children in rural and intermediate communities consistently had higher prevalence of infection across all pathogen groups compared to the urban site. These communities also had differences in household environmental conditions (Figure 2), reinforcing the importance of environmental and geographic-specific conditions in determining infection patterns. In a previous analysis, we found that WASH and animal exposure were associated with enteric pathogen detection in children aged 6 months in the ECoMiD cohort(25). The difference in enteric pathogen prevalence we observed between community types also aligns with results from a study in Guatemala, which found increased likelihood of EAEC, EPEC, ETEC, *Campylobacter,* EIEC/*Shigella*, *Giardia*, and STEC in a rural site compared to urban site(32). Additionally, in previous studies in Northern Ecuador, there was an increase likelihood of diarrhea caused by *E. coli* infections in travelers to urban sites from rural sites(33) and differences in *E. coli* pathotypes across urban and rural communities(34–36), highlighting the variability of enteric pathogen transmission within specific geographies. Our results add to multi-country studies like MAL-ED (Malnutrition and Enteric Disease multicenter longitudinal cohort study) that were not able to isolate the effects of urbanicity given that rural and urban comparisons were also between-country comparisons(37). In this study, parasites were the least prevalent enteric pathogen type overall but were notably more prevalent in the rural sites. *Giardia* and *Cryptosporidium* were the dominant parasites, similar to other studies of young children in LMICs(6,38), while the prevalence of *Ascaris* was lower than results from a systematic review that found a pooled prevalence for all ages in South America (11.58%) and for all ages in Ecuador (39.6%)(39). Despite the lower prevalence compared to previous studies, *Ascaris* infections were more common in rural communities compared to our urban community, consistent with its environmental persistence.

The prevalence of bacterial and parasitic pathogens generally increased steadily with age, with some exceptions, consistent with longitudinal findings from MAL-ED and other studies that have employed TAC for multisite comparisons of pathogen burden(5,32) and previous studies in this region(35). For example, the trends seen in this study aligned closely with the increase seen in *Shigella* prevalence (<5% at 3 months to ∼20% at 24 months), *Campylobacter* prevalence (∼10% at 3 months to ∼30% at 12 months), and *Giardia* prevalence (<10% at 3 months to >50% at 24 months) in MAL-ED(5). In contrast EAEC detection in MAL-ED peaked at 6 months then decreased and aEPEC, tEPEC, ETEC, and STEC increased with age and then plateaued between 6-12 months. The increased prevalence of infection with DAEC, EHEC, EIEC/*Shigella*, ETEC, aEPEC, *Campylobacter*, *Ascaris*, *Cryptosporidium,* and *Giardia* at ages 12 and 18 months likely reflect children’s greater environmental exploration, declining maternal antibody protection, and dietary transitions that heighten risk of enteric pathogen exposure(40–43).

*Salmonella* prevalence decreased with age, which was not observed in MAL-ED (consistently no to low detection)(5) nor in a study from Guatemala (no detection at 6-12 months [rural and urban] and 25-36 months [urban] and <10% prevalence at 13-24 months [rural and urban] and 25-36 months [rural])(32). In contrast, viral infections showed a more complex pattern: some viruses (e.g., adenovirus) increased with age, while others (e.g., enterovirus, rotavirus) declined. These divergent patterns in virus prevalence were also observed in MAL-ED (where adenovirus and sapovirus initially increased then plateaued or slightly decreased; norovirus prevalence was variable depending on genotype; and rotavirus decreased with age(5)). Virus-specific immunity, seasonal circulation, and vaccination programs (especially for rotavirus) may influence prevalence trends(4,44,45). The substantial burden of *Cryptosporidium* among 18-month-olds is concerning given its association with growth faltering and mortality in young children(46–48).

### Co-infections

The frequency and consistency of co-infections in this study was greater than seen in previous studies(4,6–8,11,32,49–55). Our rates of detection of co-infections was greater than those reported in MAL-ED, which reported co-infections in 41% of diarrheal stools and 29% of non-diarrheal stools(37); the Global Enteric Multicenter Study (GEMS), which reported co-infections in ∼70% of children with diarrhea(4); the Maputo Sanitation (MapSan) Trial, which reported co-infections in 51-66% of children depending on study arm and age(7); and the Safe Start cluster-randomized control trial, which reported co-infections in 66.9% of infants(55). This highlights that enteric co-infections remain a persistent and substantial burden, despite ongoing public health and WASH interventions. However, each of these studies assayed for a different set of pathogens, which limits the comparability of infection rates. For example, in our study, rates of co-infection were 79% when EAEC, the most commonly detected pathogen, was not included and 88% when all pathogens were included. While the prevalence of co-infections in this study was higher than previously seen, the number of pathogens per stool ranged from 0-11 with a median of 4 pathogens in this study, aligning with results from MAL-ED (range: 0-11)(6), GEMS (range: 0-7)(4), the WASH Benefits Bangladesh study (average: 3.8)(8), and a cross-sectional study conducted in Lao People’s Democratic Republic (median: 4)(56). This suggests that enteric pathogen exposure in this study is widespread and persistent, resulting in a stable background burden of co-infections, rather than being driven by a smaller subset of children with no pathogens detected or an extreme pathogen burden.

The frequent positive co-occurrence between enteric pathogens seen in this study (Figure 5) suggests a possible ecological or mechanistic linkage such as a shared transmission route(14,50). However, their co-occurrence also raises the possibility of microbial synergy or immune modulation that increases the likelihood of acquiring additional infections(57). If a negative co-occurrence was seen, this could suggest competition between the pathogens (Table S1). Bacterial pathogens dominated the co-infection profiles with a bacterial target present in 98.8% of co-infection profiles, especially EAEC, DAEC, aEPEC, and ETEC, similar to previous results(14) and likely due to the high prevalence of these pathogen as well as the higher number of bacterial pathogens assayed compared to viral and parasitic pathogens. Viral pathogens were present in 76% of co-infection profiles, with enterovirus and adenovirus frequently appearing. These pathogens co-occurred more frequently than expected by chance. While frequent co-infections and co-occurrence associations were seen, rotavirus was not one of the most frequent enteric pathogens in co-infection profiles and no positive or negative co-occurrence associations were seen, in contrasts with previous studies(50–52). Positive co-occurrence associations were seen between astrovirus and adenovirus, matching results from a study in southwest China(58). The positive co-occurrence associations between *Trichuris trichiura* with ETEC and EIEC/*Shigella* are similar to those previously seen in Laos with significant concurrent *Trichuris trichiura* and any bacterial infection(59). These findings reinforce the observation that although children often carry a diverse range of pathogens, specific pathogen groupings are more likely to appear together than by chance.

The clinical relevance of co-infection is increasingly recognized, including in asymptomatic infections(14). Studies have shown that co-infections may prolong pathogen shedding, reduce vaccine efficacy, and increase susceptibility to subsequent infections(15,60,61). Importantly, some of the most frequently co-occurring organisms—EAEC, ETEC, and *Giardia lamblia*—are implicated in long-term gut damage and environmental enteric dysfunction (EED)(62,63). This raises concern that children in these settings are continuously exposed to a mix of pathogens capable of inducing chronic inflammation and malabsorption. These conditions are difficult to detect and treat clinically. This also highlights that reducing enteric pathogen infections individually through vaccination may be insufficient if overall environmental contamination is not addressed.

### Strengths and limitations

This study utilized data from ECoMiD, a well-characterized birth cohort spanning an urban-rural gradient, enabling examination of how urbanicity, environmental conditions, and age impact enteric pathogen infections. The use of the multiplex TAC assay allowed simultaneous, sensitive detection of a broad range of bacterial, viral, and parasitic enteric pathogens, enabling analysis of co-infections, though diarrheal measurements and symptomatic illnesses are not presented due to concerns about reporting biases(64). Given the high sensitivity of this method, we may be detecting transient or non-clinically relevant pathogen carriage events. Nonetheless, high burdens of enteric co-infection detected by qPCR-based methods have been associated with long term negative health outcomes (65,66). Additionally, while molecular detection methods like TAC cannot differentiate an active infection from prolonged shedding or subclinical carriage, even subclinical infections may result in development of environmental enteric dysfunction. The small number of children in the rural river communities limited our ability to assess differences in the prevalence of infection compared to other community types, resulting in wider confidence intervals for these estimates. Repeated sampling from the same children across ages strengthened the ability to assess age-related trends in infection burden and patterns.

## Conclusions

This study adds to the growing body of evidence that high rates of enteric pathogen co-infections continue to be the norm rather than the exception for infants and young children living in low-resource settings. These findings emphasize the importance of WASH public health interventions that aim to reduce overall microbial exposure, to complement interventions such as vaccines that focus on individual pathogens. With the high prevalence of co-infections, future research should continue to explore the mechanistic consequences of co-infection on gut health, immunity, and child development.

## Data Availability

All data produced in the present study are available upon reasonable request to the authors and given IRB approval.

## ACKNOWLEDGEMENTS

We gratefully acknowledge the families who enrolled in and provided specimens for the ECoMiD study. This project would not have been possible without our dedicated local field teams who administered the surveys and collected samples: Diana Barreiro, Mariuxi Caicedo, Grace Macias, Maria Nela Obando, Karen Caucedo, Yesica Perlaza, Yina Segura, Jorge Mejia, Juleisy Palma, Ana Rosero, Tamara Valencia, Zaida Jimenez, Sara Charcopa, Daasny Monrroy, Denise Angulo, Naira Martinez, Dayanara Wila, and Adriana Lupero. This work was funded by the National Institutes of Health (R01A137679 [KL] and R01AI162867 [BFA]). The content is solely the responsibility of the authors and does not necessarily represent the official views of the National Institutes of Health. This work was supported in part by the UW NIEHS sponsored Biostatistics, Epidemiologic and Bioinformatic Training in Environmental Health (BEBTEH) Training Grant, Grant #: NIEHS T32ES015459 (CH) and the Environmental Pathology/Toxicology Training Program (EP/T) Training Grant, Grant #: NIEHS T32ES007032 (CH).

## SUPPORTING INFORMATION CAPTIONS

**S1 Figure. Histogram of stool samples collected within 6 weeks of the target age** (dashed lines), where the target age in days for 6 month samples was 182 (mean=189, range: 163-244), for 12 months was 365 (mean=370, range: 336-446), and for 18 months was 548 (mean=558, range: 534-623) days.

**S1 Table. Pathogen classification, TaqMan Array Card gene targets, and prevalence by urbanicity and age.**

**S2 Table. Co-infection pathogen profiles**

**S3 Table. Association of age (±6 weeks) and urbanicity with pathogen infection outcomes**, adjusted for the child’s birth year and season. The reference categories were 6 months for age and urban for urbanicity.

**S2 Figure. Associations between age, urbanicity, and pathogen infection.** All estimates are adjusted for the child’s birth year and season. Age models are adjusted for urbanicity and urbanicity models are adjusted for age. The *x*-axes are a count ratio for number of pathogens, odds ratio for EAEC, and a prevalence ratio for all other outcomes. Asterisks indicate significance at *p*<0.05. Where aEPEC is atypical enteropathogenic *E. coli*, DAEC is diffusely adherent *E. coli*, EAEC is enteroaggregative *E. coli*, EHEC is enterohemorrhagic *E. coli*, EIEC is enteroinvasive *E. coli*, ETEC is enterotoxigenic *E. coli*, STEC is shigatoxigenic *E. coli*, and tEPEC is typical enteropathogenic *E. coli*.

**S3 Figure. Associations between age (±6 weeks), urbanicity, and pathogen infection.** All estimates are adjusted for the child’s birth year and season. Age models are adjusted for urbanicity and urbanicity models are adjusted for age. The *x*-axes are a count ratio for number of pathogens, and a prevalence ratio for all other outcomes. Asterisks indicate significance at *p*<0.05. Where EAEC is enteroaggregative *E. coli*.

**S4 Figure. Mean number of pathogens detected per child at 6, 12, and 18 months by community type for all samples** (not limited to ±6 weeks) with error bars representing standard error.

## Notes

### Competing Interest Statement

The authors have declared no competing interest.

### Author Declarations

The study protocol was approved by the institutional review boards of the University of Washington (IRB STUDY00014270), Emory University (IRB00101202), and the Universidad San Francisco de Quito (USFQ; 2018 022M), as well as the Ministry of Health of Ecuador (MSPCURI000253 4).

## REFERENCES

1. Kotloff KL, Nataro JP, Blackwelder WC, Nasrin D, Farag TH, Panchalingam S, et al. Burden and aetiology of diarrhoeal disease in infants and young children in developing countries (the Global Enteric Multicenter Study, GEMS): a prospective, case-control study. Lancet (London, England). 2013/05/18 ed. 2013 Jul 20;382(9888):209–22. doi:10.1016/s0140-6736(13)60844-2 PubMed PMID: 23680352.

2. Troeger C. Estimates of the global, regional, and national morbidity, mortality, and aetiologies of diarrhoea in 195 countries: a systematic analysis for the Global Burden of Disease Study 2016. Lancet Infect Dis. 2018;18:1211–28.

3. Walker CL, Rudan I, Liu L, Nair H, Theodoratou E, Bhutta ZA, et al. Global burden of childhood pneumonia and diarrhoea. Lancet (London, England). 2013/04/16 ed. 2013 Apr 20;381(9875):1405–16. Located at: 23582727. doi:10.1016/s0140-6736(13)60222-6

4. Liu J, Platts-Mills JA, Juma J, Kabir F, Nkeze J, Okoi C, et al. Use of quantitative molecular diagnostic methods to identify causes of diarrhoea in children: a reanalysis of the GEMS case-control study. Lancet. 2016 Sep 24;388(10051):1291–301. doi:10.1016/S0140-6736(16)31529-X PubMed PMID: 27673470; PubMed Central PMCID: PMC5471845.

5. Rogawski ET, Liu J, Platts-Mills JA, Kabir F, Lertsethtakarn P, Siguas M, et al. Use of quantitative molecular diagnostic methods to investigate the effect of enteropathogen infections on linear growth in children in low-resource settings: longitudinal analysis of results from the MAL-ED cohort study. The Lancet Global Health. 2018 Dec;6(12):e1319–28. doi:10.1016/S2214-109X(18)30351-6 PubMed PMID: 30287125; PubMed Central PMCID: PMC6227248.

6. Platts-Mills JA, Liu J, Rogawski ET, Kabir F, Lertsethtakarn P, Siguas M, et al. Use of quantitative molecular diagnostic methods to assess the aetiology, burden, and clinical characteristics of diarrhoea in children in low-resource settings: a reanalysis of the MAL-ED cohort study. Lancet Glob Health. 2018 Dec;6(12):e1309–18. doi:10.1016/S2214-109X(18)30349-8 PubMed PMID: 30287127; PubMed Central PMCID: PMC6227251.

7. Knee J, Sumner T, Adriano Z, Anderson C, Bush F, Capone D, et al. Effects of an urban sanitation intervention on childhood enteric infection and diarrhea in Maputo, Mozambique: A controlled before-and-after trial. eLife. 2021 Apr 9;10:e62278. doi:10.7554/eLife.62278 PubMed PMID: 33835026; PubMed Central PMCID: PMC8121544.

8. Grembi JA, Lin A, Karim MA, Islam MO, Miah R, Arnold BF, et al. Effect of water, sanitation, handwashing and nutrition interventions on enteropathogens in children 14 months old: a cluster-randomized controlled trial in rural Bangladesh. J Infect Dis. 2020 Aug 29;227(3):434–47. doi:10.1093/infdis/jiaa549 PubMed PMID: 32861214; PubMed Central PMCID: PMC9891429.

9. Nataro JP, Guerrant RL. Chronic consequences on human health induced by microbial pathogens: Growth faltering among children in developing countries. Vaccine. 2017;35(49):6807–12. doi:10.1016/j.vaccine.2017.05.035

10. Amin MA, Akhtar M, Khan ZH, Islam MT, Firoj MG, Begum YA, et al. Coinfection and clinical impact of enterotoxigenic Escherichia coli harboring diverse toxin variants and colonization factors: 2017-2022. Int J Infect Dis. 2025 Feb;151:107365. doi:10.1016/j.ijid.2024.107365 PubMed PMID: 39694230; PubMed Central PMCID: PMC11798591.

11. Zhang SX, Zhou YM, Xu W, Tian LG, Chen JX, Chen SH, et al. Impact of co-infections with enteric pathogens on children suffering from acute diarrhea in southwest China. Infect Dis Poverty. 2016 Jun 27;5:64. doi:10.1186/s40249-016-0157-2 PubMed PMID: 27349521; PubMed Central PMCID: PMC4922062.

12. Praharaj I, Platts-Mills JA, Taneja S, Antony K, Yuhas K, Flores J, et al. Diarrheal Etiology and Impact of Coinfections on Rotavirus Vaccine Efficacy Estimates in a Clinical Trial of a Monovalent Human–Bovine (116E) Oral Rotavirus Vaccine, Rotavac, India. Clin Infect Dis. 2019 Jul 15;69(2):243–50. doi:10.1093/cid/ciy896 PubMed PMID: 30335135; PubMed Central PMCID: PMC6603264.

13. Potgieter N, Heine L, Ngandu JPK, Ledwaba SE, Zitha T, Mudau LS, et al. High Burden of Co-Infection with Multiple Enteric Pathogens in Children Suffering with Diarrhoea from Rural and Peri-Urban Communities in South Africa. Pathogens. 2023 Feb 14;12(2):315. doi:10.3390/pathogens12020315 PubMed PMID: 36839587; PubMed Central PMCID: PMC9959912.

14. Colgate ER, Klopfer C, Dickson DM, Lee B, Wargo MJ, Alam A, et al. Network analysis of patterns and relevance of enteric pathogen co-infections among infants in a diarrhea-endemic setting. PLOS Computational Biology. 2023 Nov 22;19(11):e1011624. doi:10.1371/journal.pcbi.1011624

15. Nembot Fogang BA, Debrah LB, Owusu M, Agyei G, Meyer J, Gmanyami JM, et al. Helminth Coinfections Modulate Disease Dynamics and Vaccination Success in the Era of Emerging Infectious Diseases. Vaccines (Basel). 2025 Apr 22;13(5):436. doi:10.3390/vaccines13050436 PubMed PMID: 40432048; PubMed Central PMCID: PMC12116102.

16. Alirol E, Getaz L, Stoll B, Chappuis F, Loutan L. Urbanisation and infectious diseases in a globalised world. The Lancet Infectious Diseases. 2011 Feb;11(2):131–41. doi:10.1016/S1473-3099(10)70223-1

17. Boyce MR, Katz R, Standley CJ. Risk Factors for Infectious Diseases in Urban Environments of Sub-Saharan Africa: A Systematic Review and Critical Appraisal of Evidence. Trop Med Infect Dis. 2019 Sep 29;4(4):123. doi:10.3390/tropicalmed4040123 PubMed PMID: 31569517; PubMed Central PMCID: PMC6958454.

18. Smith SM, Montero L, Paez M, Ortega E, Hall E, Bohnert K, et al. Locals get travellers’ diarrhoea too: Risk factors for diarrhoeal illness and pathogenic Escherichia coli infection across an urban-rural gradient in Ecuador. Trop Med Int Health. 2019 Feb;24(2):205–19. doi:10.1111/tmi.13183 PubMed PMID: 30444557; PubMed Central PMCID: PMC7476357.

19. Qadri F, Saha A, Ahmed T, Al Tarique A, Begum YA, Svennerholm AM. Disease Burden Due to Enterotoxigenic Escherichia coli in the First 2 Years of Life in an Urban Community in Bangladesh. Infection and Immunity. 2007 Aug;75(8):3961–8. doi:10.1128/iai.00459-07

20. Kraay ANM, Trostle J, Brouwer AF, Trujillo WC, Eisenberg JNS. Determinants of short-term movement in a developing region and implications for disease transmission. Epidemiology. 2018 Jan;29(1):117–25. doi:10.1097/EDE.0000000000000751 PubMed PMID: 28901976; PubMed Central PMCID: PMC5718955.

21. Eisenberg JNS, Cevallos W, Ponce K, Levy K, Bates SJ, Scott JC, et al. Environmental change and infectious disease: How new roads affect the transmission of diarrheal pathogens in rural Ecuador. PNAS. 2006 Dec 19;103(51):19460–5. doi:10.1073/pnas.0609431104 PubMed PMID: 17158216; PubMed Central PMCID: PMC1693477.

22. Freeman MC, Stocks ME, Cumming O, Jeandron A, Higgins JPT, Wolf J, et al. Systematic review: Hygiene and health: systematic review of handwashing practices worldwide and update of health effects. Trop Med Int Health. 2014 Aug;19(8):906–16. doi:10.1111/tmi.12339

23. Tanaka M, Nakayama J. Development of the gut microbiota in infancy and its impact on health in later life. Allergology International. 2017 Oct 1;66(4):515–22. doi:10.1016/j.alit.2017.07.010

24. Lee GO, Eisenberg JNS, Uruchima J, Vasco G, Smith SM, Van Engen A, et al. Gut microbiome, enteric infections and child growth across a rural-urban gradient: protocol for the ECoMiD prospective cohort study. BMJ Open. 2021 Oct 22;11(10):e046241. doi:10.1136/bmjopen-2020-046241 PubMed PMID: 34686548; PubMed Central PMCID: PMC8543627.

25. Jesser KJ, Zhou NA, Hemlock C, Miller-Petrie MK, Contreras JD, Ballard A, et al. Environmental Exposures Associated with Enteropathogen Infection in Six-Month-Old Children Enrolled in the ECoMiD Cohort along a Rural-Urban Gradient in Northern Ecuador†. Environ Sci Technol. 2025 Jan 14;59(1):103–18. doi:10.1021/acs.est.4c07753 PubMed PMID: 39807583; PubMed Central PMCID: PMC11740902.

26. Freeman M, Victor C, Garn J, Kann R, Fagnant-Sperati C, Kowalsky E, et al. The Impact of Urban Water Supply Improvements on Infant Enteric Pathogen Infection, Diarrhea, and Growth: Results from the PAASIM Matched Cohort Study [Internet]. Research Square; 2025 [cited 2026 Apr 15]. Available from: researchsquare.com/article/rs-6697339/v1 doi:10.21203/rs.3.rs-6697339/v1

27. Jesser KJ, Levy K. Updates on defining and detecting diarrheagenic Escherichia coli pathotypes. Current Opinion in Infectious Diseases. 2020 Oct;33(5):372–80. doi:10.1097/QCO.0000000000000665 PubMed PMID: 32773499; PubMed Central PMCID: PMC7819864.

28. Enteroviruses [Internet]. 2010 [cited 2026 Jun 23]. Available from: https://www.ecdc.europa.eu/en/enteroviruses

29. Capone D, Bakare T, Barker T, Chatham AH, Clark R, Copperthwaite L, et al. Risk Factors for Enteric Pathogen Exposure among Children in Black Belt Region of Alabama, USA - Volume 29, Number 12—December 2023 - Emerging Infectious Diseases journal - CDC [Internet]. doi:10.3201/eid2912.230780

30. Cannon JL, Baker JM, Mattison CP, Browne H, Nguyen K, Burke RM, et al. Enteric Viral Pathogens in Infants in the United States: 2017–2020. Pediatrics. 2026 Apr 9;157(5):e2025072461. doi:10.1542/peds.2025-072461

31. GBD 2021 Diarrhoeal Diseases Collaborators. Global, regional, and national age-sex-specific burden of diarrhoeal diseases, their risk factors, and aetiologies, 1990-2021, for 204 countries and territories: a systematic analysis for the Global Burden of Disease Study 2021. Lancet Infect Dis. 2025 May;25(5):519–36. doi:10.1016/S1473-3099(24)00691-1 PubMed PMID: 39708822; PubMed Central PMCID: PMC12018300.

32. Gaensbauer JT, Lamb M, Calvimontes DM, Asturias EJ, Kamidani S, Contreras-Roldan IL, et al. Identification of Enteropathogens by Multiplex PCR among Rural and Urban Guatemalan Children with Acute Diarrhea. Am J Trop Med Hyg. 2019 Sep;101(3):534–40. doi:10.4269/ajtmh.18-0962 PubMed PMID: 31392942; PubMed Central PMCID: PMC6726947.

33. Smith SM, Montero L, Paez M, Ortega E, Hall E, Bohnert K, et al. Locals get travellers’ diarrhoea too: risk factors for diarrhoeal illness and pathogenic Escherichia coli infection across an urban-rural gradient in Ecuador. Tropical Medicine & International Health. 2019;24(2):205–19. doi:10.1111/tmi.13183

34. Montero L, Smith SM, Jesser KJ, Paez M, Ortega E, Peña-Gonzalez A, et al. Distribution of Escherichia coli Pathotypes along an Urban–Rural Gradient in Ecuador. Am J Trop Med Hyg. 2023 Sep;109(3):559–67. doi:10.4269/ajtmh.23-0167 PubMed PMID: 37549901; PubMed Central PMCID: PMC10484266.

35. Vieira N, Bates SJ, Solberg OD, Ponce K, Howsmon R, Cevallos W, et al. High prevalence of enteroinvasive Escherichia coli isolated in a remote region of northern coastal Ecuador. Am J Trop Med Hyg. 2007 Mar;76(3):528–33. PubMed PMID: 17360879; PubMed Central PMCID: PMC2396511.

36. Vasco G, Trueba G, Atherton R, Calvopina M, Cevallos W, Andrade T, et al. Identifying etiological agents causing diarrhea in low income Ecuadorian communities. The American journal of tropical medicine and hygiene. 2014/07/23 ed. 2014 Sep;91(3):563–9. Located at: 25048373. doi:10.4269/ajtmh.13-0744 PubMed PMID: 25048373; PubMed Central PMCID: PMC4155560.

37. Platts-Mills JA, Babji S, Bodhidatta L, Gratz J, Haque R, Havt A, et al. Pathogen-specific burdens of community diarrhoea in developing countries: a multisite birth cohort study (MAL-ED). The Lancet Global Health. 2015 Sep;3(9):e564–75. doi:10.1016/S2214-109X(15)00151-5 PubMed PMID: 26202075; PubMed Central PMCID: PMC7328884.

38. Habib A, Andrianonimiadana L, Rakotondrainipiana M, Andriantsalama P, Randriamparany R, Randremanana RV, et al. High prevalence of intestinal parasite infestations among stunted and control children aged 2 to 5 years old in two neighborhoods of Antananarivo, Madagascar. PLOS Neglected Tropical Diseases. 2021 Apr 20;15(4):e0009333. doi:10.1371/journal.pntd.0009333

39. Holland C, Sepidarkish M, Deslyper G, Abdollahi A, Valizadeh S, Mollalo A, et al. Global prevalence of Ascaris infection in humans (2010–2021): a systematic review and meta-analysis. Infectious Diseases of Poverty. 2022 Nov 18;11(1):113. doi:10.1186/s40249-022-01038-z

40. Sockett PN, Rodgers FG. Enteric and foodborne disease in children: A review of the influence of food- and environment-related risk factors. Paediatr Child Health. 2001 Apr;6(4):203–9. doi:10.1093/pch/6.4.203 PubMed PMID: 20084237; PubMed Central PMCID: PMC2804543.

41. Hossain MJ, Saha D, Antonio M, Nasrin D, Blackwelder WC, Ikumapayi UN, et al. Cryptosporidium infection in rural Gambian children: Epidemiology and risk factors. PLoS Negl Trop Dis. 2019 Jul;13(7):e0007607. doi:10.1371/journal.pntd.0007607 PubMed PMID: 31348795; PubMed Central PMCID: PMC6685629.

42. Deblais L, Ojeda A, Brhane M, Mummed B, Hassen KA, Ahmedo BU, et al. Prevalence and Load of the Campylobacter Genus in Infants and Associated Household Contacts in Rural Eastern Ethiopia: a Longitudinal Study from the Campylobacter Genomics and Environmental Enteric Dysfunction (CAGED) Project. Appl Environ Microbiol. 2023 Jul 26;89(7):e0042423. doi:10.1128/aem.00424-23 PubMed PMID: 37310259; PubMed Central PMCID: PMC10370295.

43. Donowitz JR, Alam M, Kabir M, Ma JZ, Nazib F, Platts-Mills JA, et al. A Prospective Longitudinal Cohort to Investigate the Effects of Early Life Giardiasis on Growth and All Cause Diarrhea. Clin Infect Dis. 2016 Sep 15;63(6):792–7. doi:10.1093/cid/ciw391 PubMed PMID: 27313261; PubMed Central PMCID: PMC4996141.

44. Lee B, Damon CF, Platts-Mills JA. Pediatric acute gastroenteritis due to adenovirus 40/41 in low-and middle-income countries. Curr Opin Infect Dis. 2020 Oct;33(5):398–403. doi:10.1097/QCO.0000000000000663 PubMed PMID: 32773498; PubMed Central PMCID: PMC8286627.

45. Keita AM, Doh S, Sow SO, Powell H, Omore R, Jahangir Hossain M, et al. Prevalence, Clinical Severity, and Seasonality of Adenovirus 40/41, Astrovirus, Sapovirus, and Rotavirus Among Young Children With Moderate-to-Severe Diarrhea: Results From the Vaccine Impact on Diarrhea in Africa (VIDA) Study. Clin Infect Dis. 2023 Apr 1;76(Supplement_1):S123–31. doi:10.1093/cid/ciad060

46. Khalil IA, Troeger C, Rao PC, Blacker BF, Brown A, Brewer TG, et al. Morbidity, mortality, and long-term consequences associated with diarrhoea from Cryptosporidium infection in children younger than 5 years: a meta-analyses study. The Lancet Global Health. 2018 Jul 1;6(7):e758–68. doi:10.1016/S2214-109X(18)30283-3 PubMed PMID: 29903377.

47. Checkley W, Epstein LD, Gilman RH, Black RE, Cabrera L, Sterling CR. Effects of Cryptosporidium parvum infection in Peruvian children: growth faltering and subsequent catch-up growth. Am J Epidemiol. 1998 Sep 1;148(5):497–506. doi:10.1093/oxfordjournals.aje.a009675 PubMed PMID: 9737562.

48. Khan MA, Haque MA, Tariqujjaman Md, Faruque A, Ahmed T, Mahfuz M. Impact of asymptomatic Cryptosporidium infection on nutritional status in children under two: a multi-country cohort study. BMC Nutr. 2025 Oct 21;11:190. doi:10.1186/s40795-025-01183-2 PubMed PMID: 41121211; PubMed Central PMCID: PMC12542147.

49. Andersson M, Kabayiza JC, Elfving K, Nilsson S, Msellem MI, Mårtensson A, et al. Coinfection with Enteric Pathogens in East African Children with Acute Gastroenteritis—Associations and Interpretations. Am J Trop Med Hyg. 2018 Jun;98(6):1566–70. doi:10.4269/ajtmh.17-0473 PubMed PMID: 29692296; PubMed Central PMCID: PMC6086148.

50. Lindsay B, Ramamurthy T, Sen Gupta S, Takeda Y, Rajendran K, Nair GB, et al. Diarrheagenic pathogens in polymicrobial infections. Emerg Infect Dis. 2011 Apr;17(4):606–11. doi:10.3201/eid1704.100939 PubMed PMID: 21470448; PubMed Central PMCID: PMC3377398.

51. Bhavnani D, Goldstick JE, Cevallos W, Trueba G, Eisenberg JNS. Synergistic effects between rotavirus and coinfecting pathogens on diarrheal disease: evidence from a community-based study in northwestern Ecuador. Am J Epidemiol. 2012 Sep 1;176(5):387–95. doi:10.1093/aje/kws220 PubMed PMID: 22842722; PubMed Central PMCID: PMC3499114.

52. Román E, Wilhelmi I, Colomina J, Villar J, Luz Cilleruelo M, Nebreda V, et al. Acute viral gastroenteritis: proportion and clinical relevance of multiple infections in Spanish children. Journal of Medical Microbiology. 2003;52(5):435–40. doi:10.1099/jmm.0.05079-0

53. Bilenko N, Levy A, Dagan R, Deckelbaum RJ, El-On Y, Fraser D. Does co-infection with Giardia lamblia modulate the clinical characteristics of enteric infections in young children? Eur J Epidemiol. 2004;19(9):877–83. doi:10.1023/b:ejep.0000040533.75646.9c PubMed PMID: 15499898.

54. Mason J, Iturriza-Gomara M, O’Brien SJ, Ngwira BM, Dove W, Maiden MCJ, et al. Campylobacter Infection in Children in Malawi Is Common and Is Frequently Associated with Enteric Virus Co-Infections. PLOS ONE. 2013 Mar 26;8(3):e59663. doi:10.1371/journal.pone.0059663

55. Baker KK, Mumma JAO, Simiyu S, Sewell D, Tsai K, Anderson JD, et al. Environmental and behavioural exposure pathways associated with diarrhoea and enteric pathogen detection in 5-month-old, periurban Kenyan infants: a cross-sectional study. BMJ Open. 2022 Oct;12(10):e059878. doi:10.1136/bmjopen-2021-059878

56. Chard AN, Levy K, Baker KK, Tsai K, Chang HH, Thongpaseuth V, et al. Environmental and spatial determinants of enteric pathogen infection in rural Lao People’s Democratic Republic: A cross-sectional study. PLOS Neglected Tropical Diseases. 2020 Apr 8;14(4):e0008180. doi:10.1371/journal.pntd.0008180 PubMed PMID: 32289319; PubMed Central PMCID: PMC7170279.

57. Guerrant RL, DeBoer MD, Moore SR, Scharf RJ, Lima AAM. The impoverished gut—a triple burden of diarrhoea, stunting and chronic disease. Nat Rev Gastroenterol Hepatol. 2013 Apr;10(4):220–9. doi:10.1038/nrgastro.2012.239 PubMed PMID: 23229327; PubMed Central PMCID: PMC3617052.

58. Zhang SX, Zhou YM, Xu W, Tian LG, Chen JX, Chen SH, et al. Impact of co-infections with enteric pathogens on children suffering from acute diarrhea in southwest China. Infectious Diseases of Poverty. 2016 Jun 27;5(1):64. doi:10.1186/s40249-016-0157-2

59. Chard AN, Baker KK, Tsai K, Levy K, Sistrunk JR, Chang HH, et al. Associations between soil-transmitted helminthiasis and viral, bacterial, and protozoal enteroinfections: a cross-sectional study in rural Laos. Parasites Vectors. 2019 May 7;12(1):216. doi:10.1186/s13071-019-3471-2

60. El-Heneidy A, Grimwood K, Lambert SB, Ware RS. Interference Between Enteric Viruses and Live-Attenuated Rotavirus Vaccine Virus in a Healthy Australian Birth Cohort. J Infect Dis. 2023 Apr 4;228(7):851–6. doi:10.1093/infdis/jiad094 PubMed PMID: 37014728; PubMed Central PMCID: PMC10547457.

61. Praharaj I, Platts-Mills JA, Taneja S, Antony K, Yuhas K, Flores J, et al. Diarrheal Etiology and Impact of Coinfections on Rotavirus Vaccine Efficacy Estimates in a Clinical Trial of a Monovalent Human–Bovine (116E) Oral Rotavirus Vaccine, Rotavac, India. Clin Infect Dis. 2019 Jul 15;69(2):243–50. doi:10.1093/cid/ciy896 PubMed PMID: 30335135; PubMed Central PMCID: PMC6603264.

62. Crane RJ, Jones KDJ, Berkley JA. Environmental enteric dysfunction: An overview. Food Nutr Bull. 2015 Mar;36(1 0):S76–87. doi:10.1177/15648265150361S113 PubMed PMID: 25902619; PubMed Central PMCID: PMC4472379.

63. Fahim SM, Das S, Gazi MA, Mahfuz M, Ahmed T. Association of intestinal pathogens with faecal markers of environmental enteric dysfunction among slum-dwelling children in the first 2 years of life in Bangladesh. Trop Med Int Health. 2018 Nov;23(11):1242–50. doi:10.1111/tmi.13141 PubMed PMID: 30133067; PubMed Central PMCID: PMC6282798.

64. Watson SI, Rego RTT, Hofer T, Lilford RJ. Evaluations of water, sanitation and hygiene interventions should not use diarrhoea as (primary) outcome. BMJ Glob Health. 2022 May;7(5):e008521. doi:10.1136/bmjgh-2022-008521 PubMed PMID: 35550338; PubMed Central PMCID: PMC9109038.

65. Rogawski ET, Liu J, Platts-Mills JA, Kabir F, Lertsethtakarn P, Siguas M, et al. Use of quantitative molecular diagnostic methods to investigate the effect of enteropathogen infections on linear growth in children in low-resource settings: longitudinal analysis of results from the MAL-ED cohort study. Lancet Glob Health. 2018 Dec;6(12):e1319–28. doi:10.1016/S2214-109X(18)30351-6 PubMed PMID: 30287125; PubMed Central PMCID: PMC6227248.

66. Kabir F, Iqbal J, Jamil Z, Iqbal NT, Mallawaarachchi I, Aziz F, et al. Impact of enteropathogens on faltering growth in a resource-limited setting. Front Nutr. 2023 Jan 10;9. doi:10.3389/fnut.2022.1081833

