## Supplementary material for "Enteric pathogen burden and co-infection patterns across age and a rural-urban gradient: findings from the ECoMiD birth cohort, Northern Ecuador": S1 Figure

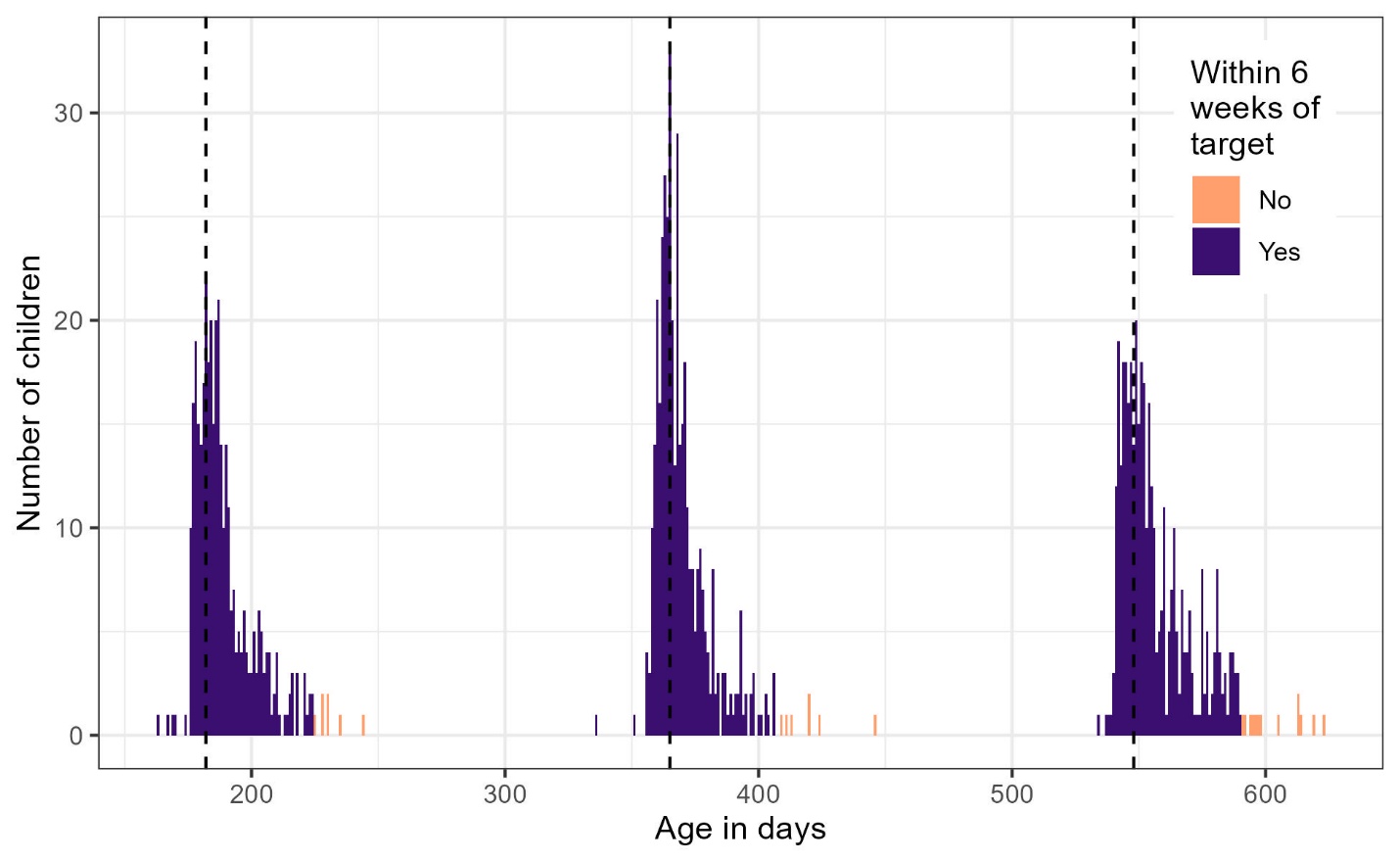

Figure S1. Histogram of stool samples collected within 6 weeks of the target age (dashed lines), where the target age in days for 6 month samples was 182 (mean=189, range: 163-244), for 12 months was 365 (mean=370, range: 336-446), and for 18 months was 548 (mean=558, range: 534-623) days.
