## Supplementary material for "Enteric pathogen burden and co-infection patterns across age and a rural-urban gradient: findings from the ECoMiD birth cohort, Northern Ecuador": S1 Table

Table S1. Pathogen classification, TaqMan Array Card gene targets, and prevalence by urbanicity and age.

| Pathogen | Targets | Overall,  n=1,154 | Urbanicity | | | |  | Age | | |
| --- | --- | --- | --- | --- | --- | --- | --- | --- | --- | --- |
|  |  |  | Urban,  n=265 | Intermediate, n=349 | Rural road,  n=420 | Rural river,  n=120 |  | 06m,  n=359 | 12m,  n=404 | 18m,  n=391 |
| Any pathogen | - | 1,102 (95%) | 243 (92%) | 339  (97%) | 405 (96%) | 115 (96%) |  | 326 (91%) | 393 (97%) | 383 (98%) |
| Any pathogen EAEC excluded | - | 1,056  (92%) | 230  (87%) | 326  (93%) | 390  (93%) | 110  (92%) |  | 298  (83%) | 382  (95%) | 376  (96%) |
| Any co-infection | - | 1,010 (88%) | 216 (82%) | 316  (91%) | 375 (89%) | 103 (86%) |  | 273 (76%) | 371 (92%) | 366 (94%) |
| Any co-infection EAEC excluded | - | 909  (79%) | 187  (71%) | 286  (82%) | 343  (82%) | 93  (78%) |  | 230  (64%) | 336  (83%) | 343  (88%) |
| Number of pathogens | - |  |  |  |  |  |  |  |  |  |
| Median (IQR) |  | 4 (2, 5) | 3 (2, 5) | 4 (3, 5) | 4 (3, 5) | 4 (3, 5) |  | 3 (2, 4) | 4 (3, 5) | 4 (3, 6) |
| Range |  | 0, 11 | 0, 9 | 0, 11 | 0, 11 | 0, 11 |  | 0, 8 | 0, 11 | 0, 11 |
| Bacteria | | | | | | | | | | |
| Any bacterial pathogen | - | 1,071 (93%) | 234 (88%) | 327  (94%) | 396 (94%) | 114 (95%) |  | 303 (84%) | 388 (96%) | 380 (97%) |
| EAEC | aaiC+, aatA+, and/or aggR+ | 842 (73%) | 178 (67%) | 262  (76%) | 306 (73%) | 96 (81%) |  | 234 (66%) | 326 (81%) | 282 (72%) |
| Missing |  | 7 | 0 | 4 | 1 | 2 |  | 4 | 0 | 3 |
| DAEC | afaB+ | 530 (46%) | 106 (40%) | 162  (46%) | 201 (48%) | 61 (51%) |  | 134 (37%) | 193 (48%) | 203 (52%) |
| STEC | eae- and stx1+ and/or stx2+ | 19 (1.6%) | 1 (0.4%) | 4  (1.1%) | 10 (2.4%) | 4  (3.3%) |  | 2 (0.6%) | 9  (2.2%) | 8  (2.0%) |
| Missing |  | 2 | 0 | 1 | 1 | 0 |  | 1 | 1 | 0 |
| aEPEC | bfpA- and eae+ | 509 (44%) | 118 (45%) | 161  (46%) | 183 (44%) | 47 (39%) |  | 99 (28%) | 206 (51%) | 204 (52%) |
| Missing |  | 2 | 0 | 1 | 1 | 0 |  | 1 | 1 | 0 |
| tEPEC | bfpA+ and eae+ | 157 (14%) | 33 (12%) | 49  (14%) | 55 (13%) | 20 (17%) |  | 36 (10%) | 70 (17%) | 51 (13%) |
| Missing |  | 2 | 0 | 1 | 1 | 0 |  | 1 | 1 | 0 |
| EHEC | eae+ and stx1+ and/or stx2+ | 117 (10%) | 22 (8.3%) | 40  (11%) | 46 (11%) | 9  (7.5%) |  | 8 (2.2%) | 49 (12%) | 60 (15%) |
| Missing |  | 2 | 0 | 1 | 1 | 0 |  | 1 | 1 | 0 |
| ETEC | LT+, STh+, and/or STp+ | 366 (32%) | 71 (27%) | 112  (32%) | 136 (32%) | 47 (40%) |  | 58 (16%) | 143 (35%) | 165 (42%) |
| Missing |  | 4 | 1 | 1 | 0 | 2 |  | 4 | 0 | 0 |
| EIEC or *Shigella* | ipaH+ and/or virF+ | 145 (13%) | 20 (7.5%) | 48  (14%) | 58 (14%) | 19 (16%) |  | 16 (4.5%) | 46 (11%) | 83 (21%) |
| *Campylobacter* spp. | cadF+, hipO+, and/or GlyA+ | 240 (21%) | 53 (20%) | 68  (19%) | 96 (23%) | 23 (19%) |  | 48 (13%) | 104 (26%) | 88 (23%) |
| *Salmonella enterica* | ttr+, tviB+, and/or STY0201+ | 76 (6.6%) | 21 (7.9%) | 25  (7.2%) | 26 (6.2%) | 4  (3.3%) |  | 37 (10%) | 27 (6.7%) | 12 (3.1%) |
| Viruses | | | | | | | | | | |
| Any viral pathogen | - | 705 (61%) | 134 (51%) | 225  (64%) | 273 (65%) | 73 (61%) |  | 216 (60%) | 255 (63%) | 234 (60%) |
| Adenovirus | Hexon+ and/or fiber gene+ | 359 (32%) | 64 (25%) | 109  (32%) | 146 (35%) | 40 (33%) |  | 74 (21%) | 134 (34%) | 151 (39%) |
| Missing |  | 23 | 7 | 10 | 6 | 0 |  | 4 | 11 | 8 |
| Astrovirus | Capsid | 61 (5.3%) | 10 (3.8%) | 16  (4.6%) | 29 (6.9%) | 6  (5.0%) |  | 15 (4.2%) | 29 (7.2%) | 17 (4.3%) |
| Enterovirus | 5’ UTR | 416 (36%) | 72 (27%) | 133  (38%) | 169 (40%) | 42 (35%) |  | 147 (41%) | 143 (35%) | 126 (32%) |
| Norovirus | GI ORF1-ORF2+ and/or GII ORF1-ORF2+ | 122 (11%) | 21 (7.9%) | 37  (11%) | 48 (11%) | 16 (13%) |  | 31 (8.6%) | 48 (12%) | 43 (11%) |
| Rotavirus | NSP3 | 38 (3.3%) | 6 (2.3%) | 12  (3.4%) | 14 (3.3%) | 6  (5.0%) |  | 18 (5.0%) | 9  (2.2%) | 11 (2.8%) |
| Sapovirus | RdRp | 40 (3.5%) | 7 (2.6%) | 16  (4.6%) | 11 (2.6%) | 6  (5.0%) |  | 9 (2.5%) | 19 (4.7%) | 12 (3.1%) |
| Parasites | | | | | | | | | | |
| Any parasitic pathogen | - | 346 (30%) | 56 (21%) | 121  (35%) | 131 (31%) | 38 (32%) |  | 37 (10%) | 114 (28%) | 195 (50%) |
| *Ascaris lumbricoides* | ITS1 | 91 (7.9%) | 4 (1.5%) | 35  (10%) | 39 (9.3%) | 13 (11%) |  | 6 (1.7%) | 27 (6.7%) | 58 (15%) |
| *Cryptosporidium* spp. | 18S+, C. hominus LIB13+, and/or C. parvum LIB 13+ | 99 (8.6%) | 17 (6.4%) | 31  (8.9%) | 35 (8.3%) | 16 (13%) |  | 12 (3.3%) | 34 (8.4%) | 53 (14%) |
| *Cyclospora cayetanensis* | 18S | 3  (0.3%) | 1 (0.4%) | 1  (0.3%) | 0  (0%) | 1  (0.8%) |  | 1 (0.3%) | 0  (0%) | 2  (0.5%) |
| *Entamoeba histolytica* | 18S | 3  (0.3%) | 0 (0%) | 1  (0.3%) | 2  (0.5%) | 0  (0%) |  | 1 (0.3%) | 1  (0.2%) | 1  (0.3%) |
| *Giardia lamblia* | 18S | 214 (19%) | 46 (17%) | 79  (23%) | 71 (17%) | 18 (15%) |  | 19 (5.3%) | 67 (17%) | 128 (33%) |
| *Trichuris trichiura* | 18S | 21 (1.8%) | 1 (0.4%) | 5  (1.4%) | 14 (3.3%) | 1  (0.8%) |  | 3 (0.8%) | 8  (2.0%) | 10 (2.6%) |
