## Supplementary material for "Enteric pathogen burden and co-infection patterns across age and a rural-urban gradient: findings from the ECoMiD birth cohort, Northern Ecuador": S2 Figure

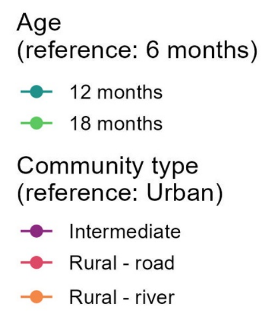

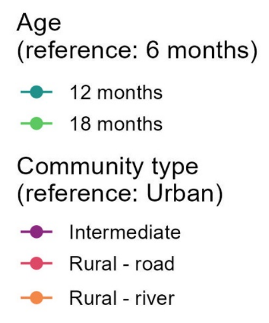

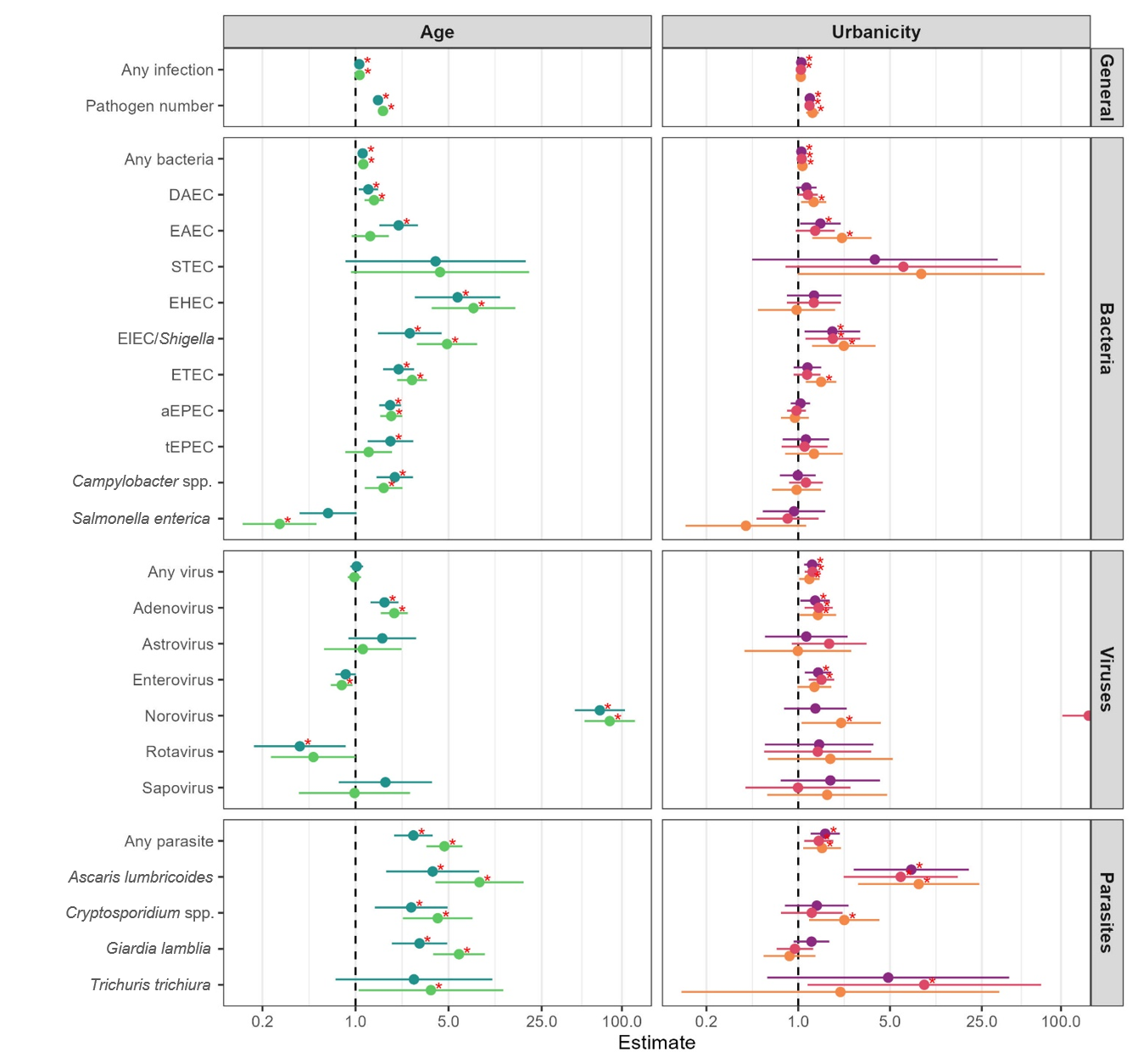

Figure S2. Associations between age, urbanicity, and pathogen infection. All estimates are adjusted for the child’s birth year and season. Age models are adjusted for urbanicity and urbanicity models are adjusted for age. The *x*-axes are a count ratio for number of pathogens, odds ratio for EAEC, and a prevalence ratio for all other outcomes. Asterisks indicate significance at *p*<0.05. Where aEPEC is atypical enteropathogenic *E. coli*, DAEC is diffusely adherent *E. coli*, EAEC is enteroaggregative *E. coli*, EHEC is enterohemorrhagic *E. coli*, EIEC is enteroinvasive *E. coli*, ETEC is enterotoxigenic *E. coli*, STEC is shigatoxigenic *E. coli*, and tEPEC is typical enteropathogenic *E. coli*.
