## Supplementary material for "Enteric pathogen burden and co-infection patterns across age and a rural-urban gradient: findings from the ECoMiD birth cohort, Northern Ecuador": S3 Figure

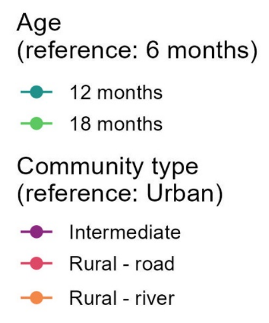

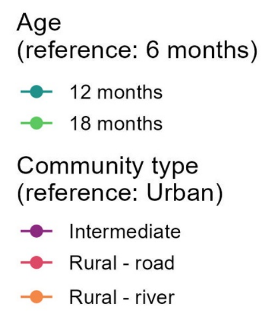

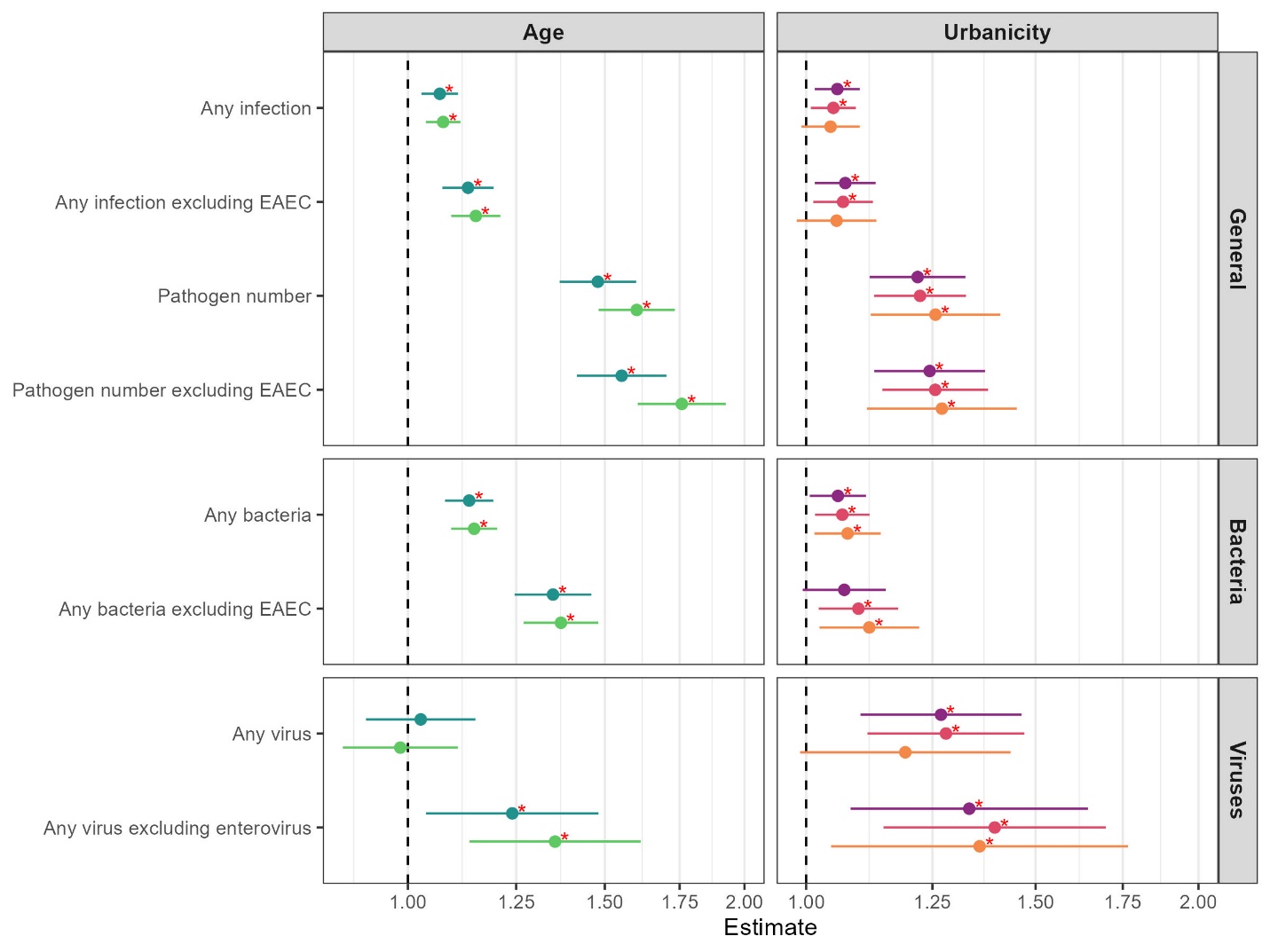

Figure S3. Associations between age (±6 weeks), urbanicity, and pathogen infection. All estimates are adjusted for the child’s birth year and season. Age models are adjusted for urbanicity and urbanicity models are adjusted for age. The *x*-axes are a count ratio for number of pathogens, and a prevalence ratio for all other outcomes. Asterisks indicate significance at *p*<0.05. Where EAEC is enteroaggregative *E. coli.*
