## Supplementary material for "Enteric pathogen burden and co-infection patterns across age and a rural-urban gradient: findings from the ECoMiD birth cohort, Northern Ecuador": S3 Table

Table S3. Association of age (±6 weeks) and urbanicity with pathogen infection outcomes, adjusted for the child’s birth year and season. The reference categories were 6 months for age and urban for urbanicity.

| Outcome | Exposure^1^ | Level | Prevalence Ratio | *p*-value |
| --- | --- | --- | --- | --- |
| Any infection | Age | 12 months | 1.07 (1.03,  1.11) | <0.001 |
|  |  | 18 months | 1.08 (1.04,  1.11) | <0.001 |
|  | Urbanicity | Intermediate | 1.06 (1.02,  1.10) | 0.00647 |
|  |  | Rural road | 1.05 (1.01,  1.09) | 0.0176 |
|  |  | Rural river | 1.04 (0.99,  1.10) | 0.1 |
| Any infection excluding EAEC | Age | 12 months | 1.13 (1.07,  1.19) | <0.001 |
|  |  | 18 months | 1.15 (1.09,  1.21) | <0.001 |
|  | Urbanicity | Intermediate | 1.07 (1.02,  1.13) | 0.0114 |
|  |  | Rural road | 1.07 (1.01,  1.13) | 0.0150 |
|  |  | Rural river | 1.06 (0.98,  1.13) | 0.134 |
| Pathogen number | Age | 12 months | 1.48 (1.37,  1.60) ^2^ | <0.001 |
|  |  | 18 months | 1.60 (1.48,  1.73) ^2^ | <0.001 |
|  | Urbanicity | Intermediate | 1.22 (1.12,  1.33) ^2^ | <0.001 |
|  |  | Rural road | 1.22 (1.13,  1.33) ^2^ | <0.001 |
|  |  | Rural river | 1.26 (1.12,  1.41) ^2^ | <0.001 |
| Pathogen number excluding EAEC | Age | 12 months | 1.55 (1.42,  1.70) ^2^ | <0.001 |
|  |  | 18 months | 1.76 (1.61,  1.92) ^2^ | <0.001 |
|  | Urbanicity | Intermediate | 1.24 (1.13,  1.37) ^2^ | <0.001 |
|  |  | Rural road | 1.26 (1.14,  1.38) ^2^ | <0.001 |
|  |  | Rural river | 1.27 (1.11,  1.45) ^2^ | <0.001 |
| Bacteria | | | | |
| Any bacteria | Age | 12 months | 1.13 (1.08,  1.19) | <0.001 |
|  |  | 18 months | 1.15 (1.09,  1.20) | <0.001 |
|  | Urbanicity | Intermediate | 1.06 (1.01,  1.11) | 0.0266 |
|  |  | Rural road | 1.07 (1.02,  1.12) | 0.00916 |
|  |  | Rural river | 1.08 (1.01,  1.14) | 0.0144 |
| Any bacteria excluding EAEC | Age | 12 months | 1.35 (1.25,  1.46) | <0.001 |
|  |  | 18 months | 1.37 (1.27,  1.48) | <0.001 |
|  | Urbanicity | Intermediate | 1.07 (0.99,  1.15) | 0.0718 |
|  |  | Rural road | 1.10 (1.02,  1.18) | 0.00975 |
|  |  | Rural river | 1.12 (1.02,  1.22) | 0.0129 |
| DAEC | Age | 12 months | 1.24 (1.05,  1.47) | 0.0125 |
|  |  | 18 months | 1.35 (1.14,  1.60) | <0.001 |
|  | Urbanicity | Intermediate | 1.14 (0.95,  1.37) | 0.146 |
|  |  | Rural road | 1.19 (1.00,  1.42) | 0.0511 |
|  |  | Rural river | 1.28 (1.02,  1.61) | 0.0333 |
| EAEC | Age | 12 months | 2.12 (1.51,  2.97) ^3^ | <0.001 |
|  |  | 18 months | 1.28 (0.93,  1.77) ^3^ | 0.131 |
|  | Urbanicity | Intermediate | 1.55 (1.08,  2.22) ^3^ | 0.0171 |
|  |  | Rural road | 1.37 (0.97,  1.93) ^3^ | 0.0707 |
|  |  | Rural river | 2.24 (1.31,  3.83) ^3^ | 0.00334 |
| STEC | Age | 12 months | 4.08 (0.87, 19.2) | 0.0751 |
|  |  | 18 months | 4.07 (0.86, 19.4) | 0.0778 |
|  | Urbanicity | Intermediate | 3.10 (0.34, 28.0) | 0.313 |
|  |  | Rural road | 6.05 (0.77, 47.6) | 0.0871 |
|  |  | Rural river | 9.08 (1.05, 78.3) | 0.0448 |
| EHEC | Age | 12 months | 5.83 (2.79, 12.2) | <0.001 |
|  |  | 18 months | 7.43 (3.60, 15.4) | <0.001 |
|  | Urbanicity | Intermediate | 1.39 (0.86,  2.26) | 0.182 |
|  |  | Rural road | 1.37 (0.85,  2.23) | 0.2 |
|  |  | Rural river | 0.90 (0.43,  1.88) | 0.776 |
| EIEC or *Shigella* | Age | 12 months | 2.48 (1.43,  4.30) | 0.00119 |
|  |  | 18 months | 4.74 (2.81,  7.99) | <0.001 |
|  | Urbanicity | Intermediate | 1.79 (1.10,  2.92) | 0.0194 |
|  |  | Rural road | 1.82 (1.13,  2.95) | 0.0141 |
|  |  | Rural river | 2.08 (1.17,  3.71) | 0.0127 |
| ETEC | Age | 12 months | 2.16 (1.65,  2.83) | <0.001 |
|  |  | 18 months | 2.64 (2.03,  3.44) | <0.001 |
|  | Urbanicity | Intermediate | 1.19 (0.93,  1.52) | 0.169 |
|  |  | Rural road | 1.18 (0.93,  1.50) | 0.166 |
|  |  | Rural river | 1.46 (1.10,  1.94) | 0.0088 |
| aEPEC | Age | 12 months | 1.85 (1.53,  2.25) | <0.001 |
|  |  | 18 months | 1.89 (1.56,  2.29) | <0.001 |
|  | Urbanicity | Intermediate | 1.04 (0.87,  1.23) | 0.683 |
|  |  | Rural road | 0.98 (0.83,  1.15) | 0.768 |
|  |  | Rural river | 0.89 (0.68,  1.15) | 0.368 |
| tEPEC | Age | 12 months | 1.81 (1.22,  2.69) | 0.00304 |
|  |  | 18 months | 1.26 (0.84,  1.89) | 0.268 |
|  | Urbanicity | Intermediate | 1.13 (0.75,  1.70) | 0.553 |
|  |  | Rural road | 1.09 (0.73,  1.63) | 0.685 |
|  |  | Rural river | 1.38 (0.84,  2.29) | 0.205 |
| *Campylobacter* spp. | Age | 12 months | 1.89 (1.38,  2.59) | <0.001 |
|  |  | 18 months | 1.64 (1.19,  2.27) | 0.00265 |
|  | Urbanicity | Intermediate | 0.96 (0.70,  1.32) | 0.808 |
|  |  | Rural road | 1.14 (0.85,  1.53) | 0.386 |
|  |  | Rural river | 0.98 (0.64,  1.52) | 0.933 |
| *Salmonella enterica* | Age | 12 months | 0.61 (0.37,  1.01) | 0.0552 |
|  |  | 18 months | 0.28 (0.15,  0.53) | <0.001 |
|  | Urbanicity | Intermediate | 0.90 (0.52,  1.56) | 0.703 |
|  |  | Rural road | 0.79 (0.46,  1.37) | 0.406 |
|  |  | Rural river | 0.41 (0.14,  1.19) | 0.102 |
| Viruses | | | | |
| Any virus | Age | 12 months | 1.03 (0.92,  1.15) | 0.64 |
|  |  | 18 months | 0.98 (0.87,  1.11) | 0.798 |
|  | Urbanicity | Intermediate | 1.27 (1.10,  1.46) | 0.00102 |
|  |  | Rural road | 1.28 (1.11,  1.47) | <0.001 |
|  |  | Rural river | 1.19 (0.99,  1.44) | 0.0647 |
| Any virus excluding enterovirus | Age | 12 months | 1.24 (1.04,  1.48) | 0.0177 |
|  |  | 18 months | 1.35 (1.14,  1.62) | <0.001 |
|  | Urbanicity | Intermediate | 1.33 (1.08,  1.65) | 0.00704 |
|  |  | Rural road | 1.40 (1.15,  1.70) | <0.001 |
|  |  | Rural river | 1.36 (1.05,  1.77) | 0.0220 |
| Adenovirus | Age | 12 months | 1.64 (1.29,  2.10) | <0.001 |
|  |  | 18 months | 1.92 (1.51,  2.43) | <0.001 |
|  | Urbanicity | Intermediate | 1.30 (1.00,  1.68) | 0.0501 |
|  |  | Rural road | 1.41 (1.10,  1.80) | 0.00652 |
|  |  | Rural river | 1.35 (0.97,  1.88) | 0.0731 |
| Astrovirus | Age | 12 months | 1.56 (0.87,  2.81) | 0.136 |
|  |  | 18 months | 1.10 (0.56,  2.19) | 0.779 |
|  | Urbanicity | Intermediate | 1.26 (0.59,  2.66) | 0.552 |
|  |  | Rural road | 1.77 (0.89,  3.50) | 0.103 |
|  |  | Rural river | 1.14 (0.44,  2.94) | 0.794 |
| Enterovirus | Age | 12 months | 0.84 (0.70,  1.01) | 0.0616 |
|  |  | 18 months | 0.79 (0.65,  0.95) | 0.0145 |
|  | Urbanicity | Intermediate | 1.41 (1.11,  1.78) | 0.0047 |
|  |  | Rural road | 1.50 (1.19,  1.88) | <0.001 |
|  |  | Rural river | 1.28 (0.94,  1.74) | 0.124 |
| Norovirus | Age | 12 months | 1.36 (0.80,  2.31) ^3^ | 0.256 |
|  |  | 18 months | 1.25 (0.73,  2.14) ^3^ | 0.409 |
|  | Urbanicity | Intermediate | 1.35 (0.77,  2.37) ^3^ | 0.288 |
|  |  | Rural road | 1.45 (0.77,  2.71) ^3^ | 0.252 |
|  |  | Rural river | 1.76 (0.89,  3.49) ^3^ | 0.107 |
| Rotavirus | Age | 12 months | 0.38 (0.17,  0.84) | 0.0172 |
|  |  | 18 months | 0.50 (0.24,  1.04) | 0.062 |
|  | Urbanicity | Intermediate | 1.41 (0.54,  3.67) | 0.478 |
|  |  | Rural road | 1.36 (0.53,  3.48) | 0.524 |
|  |  | Rural river | 1.80 (0.60,  5.39) | 0.295 |
| Sapovirus | Age | 12 months | 1.58 (0.70,  3.54) | 0.271 |
|  |  | 18 months | 0.98 (0.37,  2.60) | 0.976 |
|  | Urbanicity | Intermediate | 1.65 (0.68,  3.96) | 0.267 |
|  |  | Rural road | 0.97 (0.39,  2.46) | 0.956 |
|  |  | Rural river | 1.70 (0.59,  4.86) | 0.325 |
| Parasites | | | | |
| Any parasite | Age | 12 months | 2.78 (1.98,  3.91) | <0.001 |
|  |  | 18 months | 4.85 (3.52,  6.67) | <0.001 |
|  | Urbanicity | Intermediate | 1.63 (1.26,  2.12) | <0.001 |
|  |  | Rural road | 1.48 (1.14,  1.91) | 0.00303 |
|  |  | Rural river | 1.53 (1.09,  2.16) | 0.0147 |
| *Ascaris lumbricoides* | Age | 12 months | 4.24 (1.79, 10.1) | 0.00105 |
|  |  | 18 months | 9.43 (4.15, 21.4) | <0.001 |
|  | Urbanicity | Intermediate | 6.73 (2.44, 18.5) | <0.001 |
|  |  | Rural road | 5.98 (2.20, 16.3) | <0.001 |
|  |  | Rural river | 7.35 (2.47, 21.8) | <0.001 |
| *Cryptosporidium* spp. | Age | 12 months | 2.52 (1.34,  4.74) | 0.00421 |
|  |  | 18 months | 4.01 (2.19,  7.33) | <0.001 |
|  | Urbanicity | Intermediate | 1.35 (0.77,  2.37) | 0.292 |
|  |  | Rural road | 1.26 (0.74,  2.17) | 0.395 |
|  |  | Rural river | 2.13 (1.13,  4.01) | 0.0194 |
| *Giardia lamblia* | Age | 12 months | 3.12 (1.92,  5.09) | <0.001 |
|  |  | 18 months | 6.15 (3.89,  9.74) | <0.001 |
|  | Urbanicity | Intermediate | 1.29 (0.94,  1.77) | 0.114 |
|  |  | Rural road | 0.98 (0.71,  1.35) | 0.891 |
|  |  | Rural river | 0.87 (0.54,  1.41) | 0.578 |
| *Trichuris trichiura* | Age | 12 months | 2.85 (0.73, 11.2) | 0.134 |
|  |  | 18 months | 3.32 (0.95, 11.6) | 0.597 |
|  | Urbanicity | Intermediate | 4.07 (0.47, 35.0) | 0.201 |
|  |  | Rural road | 9.23 (1.19, 71.9) | 0.0338 |
|  |  | Rural river | 2.30 (0.14, 37.4) | 0.557 |

^1^Age models are adjusted for urbanicity and urbanicity models are adjusted for age.

^2^ Pathogen number model outcomes are count ratios.

^3^ EAEC and norovirus model outcomes are odds ratios.
