## Supplementary material for "Enteric pathogen burden and co-infection patterns across age and a rural-urban gradient: findings from the ECoMiD birth cohort, Northern Ecuador": S4 Figure

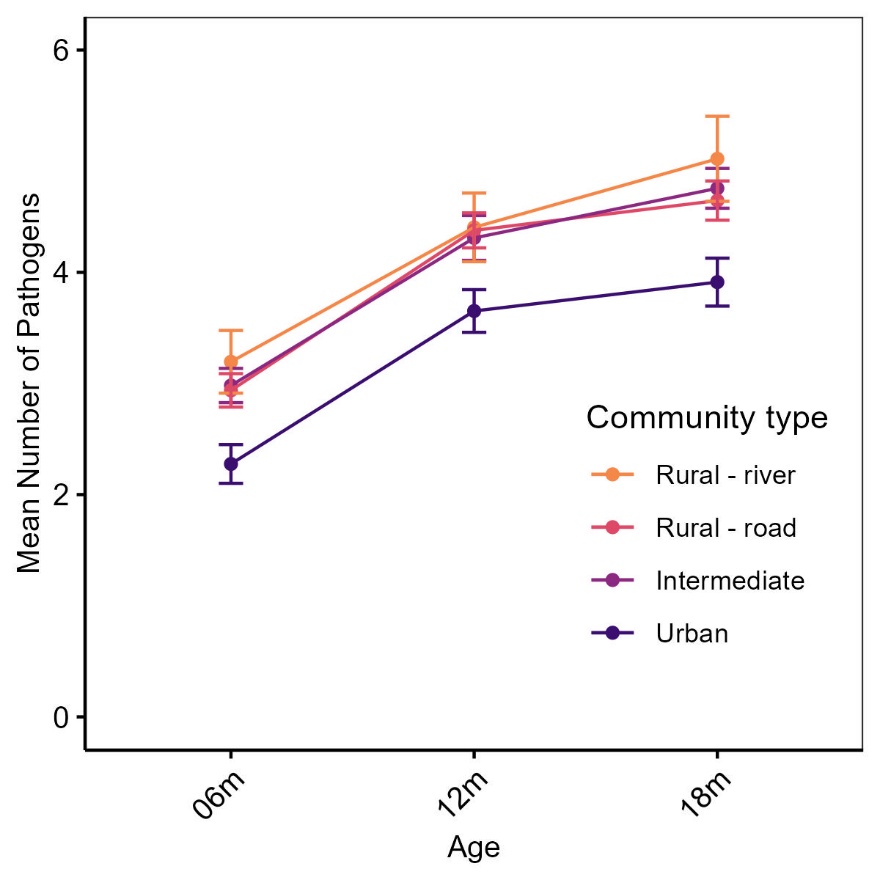
Figure S4. Mean number of pathogens detected per child at 6, 12, and 18 months by community type for all samples (not limited to ±6 weeks) with error bars representing standard error.
